# An explicit formula for minimizing the infected peak in an SIR epidemic model when using a fixed number of complete lockdowns

**DOI:** 10.1101/2021.04.11.21255289

**Authors:** Eduardo D. Sontag

## Abstract

Careful timing of NPIs (non-pharmaceutical interventions) such as social distancing may avoid high “second waves” of infections of COVID-19. This paper asks what should be the timing of a set of *k* complete-lockdowns of prespecified lengths (such as two weeks) so as to minimize the peak of the infective compartment. Perhaps surprisingly, it is possible to give an explicit and easily computable rule for when each lockdown should commence. Simulations are used to show that the rule remains fairly accurate even if lockdowns are not perfect.

## 1 Introduction

The year 2020 will be remembered for the COVID-19 (coronavirus disease 2019) pandemic, which is an individual-to-individual infection by SARS-CoV-2 (severe acute respiratory syndrome coronavirus 2). An immediate way to stop transmissibility is to establish total lockdowns, as China and Northern Italy did early on in the outbreak [1]. Lockdowns and other NPIs (non-pharmaceutical interventions) such as quarantine and social distancing were soon implemented around the world. On the other hand, frustration with isolation rules and economic costs mean that, in most countries, long lockdowns are not feasible, nor is it easy to enforce even milder forms of NPIs [2, 3, 4]. However, relaxation of NPIs could lead to “second waves” of infections when the NPIs are relaxed. This motivates the search for optimally timing the start of multiple short NPIs so as to minimize the maximum peak of infective individuals. This paper deals with that problem.

To be precise, we consider an SIR model with strict (no-contact) lockdowns, and assume that policymakers want to decide when to start each one of *K* of lockdowns, with respective lengths *T*_*k*_, *k* = 1, …, *K*. For example, the *T*_*k*_’s may all be equal, say two weeks. We provide an exact and very simple rule, which says that lockdowns should commence whenever the number of infectious individuals reaches a certain level, namely 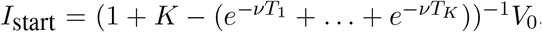, where *V*_0_ is a number that can be easily computed from the infectivity rate, recovery rate (*ν*), and the initial populations of susceptible and infected individuals. The formula for *I*_start_ simplifies in the case in which all *T*_*k*_’s are equal, *T*_*k*_ = *T*, to just (1 + *K* − *Ke*^−*νT*^)^−1^*V*_0_. In addition, we show that there will be exactly *K* peaks of infected populations, all equal to this value. Observe that as the lockdown intervals become larger, *T* →∞, the best possible maximum peak is *V*_0_*/*(1 + *K*). (Using a larger value of *T* will lengthen the duration of the pandemic but, asymptotically, not change the peak value. If there is a hope for a quick cure or vaccines, this is not necesarily a good strategy.) Obviously, a perfect or near-perfect lockdown is not practical, except in jurisdictions where complete obedience can be strongly enforced, but this theoretical problem is nonetheless of interest. Moreover, studying this case also helps understand the problem with non-strict lockdowns, as we will discuss.

### Infections and mathematical modeling

Infectious agents have critically influenced the history of mankind, with disease-causing pathogens constantly emerging or evolving. From the Plague of Athens (430-428 BC), to the fourteenth century Black Death that killed about a third of Europe’s population, to the Yellow Fever epidemic in Philadelphia in 1793, in which a tenth of the population of the city perished, to the 1918 “Spanish flu” pandemic (which did not originate in Spain) that resulted in about 3-5% of the world population dying, to the COVID-19 2020-2021 pandemic, infectious diseases have had major impacts on health, psychological and social well-being, medical advances (mRNA vaccines, for example), economics, politics, military history, and religious and racial persecution. Different types of pathogens are involved in infectious diseases. Viruses cause the common cold, influenza, measles, West Nile, and COVID-19, while anthrax, salmonella, chlamydia, and cholera are caused by bacteria, and protozoa give rise to malaria and trypanosomiasis (sleeping sickness). There are many mechanisms for transmission, including respiratory droplets (influenza, colds), body secretions (chlamydia), flies (trypanosomiasis), mosquitoes (malaria), and food or water (cholera). Control strategies include behavioral and sanitation changes (NPIs), vaccines, antibiotics, antiviral drugs. Notwithstanding this variety, there is a common mathematical structure.

The modeling of infectious diseases and their spread is an important part of mathematical biology, specifically mathematical epidemiology. Modeling is an important tool for gauging the impact of NPIs such as social distancing, masking, lock-downs, or school closings, as well as predicting/attenuating magnitude of peak infections (“flattening the curve” so as not to overwhelm ICU capacities), predicting/delaying peak infections (until vaccine/treatments available), and devising strategies for vaccination, control, or eradication of diseases. The social and political use of epidemic models must take into account their degree of realism. Good models do not incorporate all possible effects, but rather focus on the basic mechanisms in their simplest possible fashion. Not only it is difficult to model every detail, but the more details the more the likelihood of making the model sensitive to parameters and assumptions, and the more difficult it is to *understand and interpret* the model as well as to play “*what-if* “ scenarios to compare alternative containment policies. It turns out that even simple models help pose important questions about the underlying mechanisms of infection spread and possible means of control of an epidemic.

Most mathematical epidemiology models incorporate some version of the classical SIR model proposed by Kermack and McKendrick in 1927 [5]. We will restrict attention to this core model, which is suitable for describing initial stages of an infection in a single city, and also for modeling later stages when community spread becomes dominant. Mathematical models have long played a central role in epidemiology, and this has been especially true with the COVID-19 pandemic [6, 7, 8, 9, 10, 11, 12, 13, 14]. This includes control-theoretic aspects, especially optimal control [15, 16, 17, 18].

Mathematical models had a major impact on the political response to the COVID-19 pandemic. To quote from “Behind the virus report that jarred the U.S. and the U.K. to action” (*New York Times*, 17 March 2020):

> *The report [from Imperial College London], which warned that an uncontrolled spread of the disease could cause as many as 510,000 deaths in Britain, triggered a sudden shift in the government’s comparatively relaxed response to the virus. American officials said the report, which projected up to 2*.*2 million deaths in the United States from such a spread, also influenced the White House to strengthen its measures to isolate members of the public*.

Of course, one should always keep in mind as well the following quote from Dr. Anthony Fauci, Director, National Institute of Allergy and Infectious Diseases, United States (*CNN*, 05 May 2020):

> *I have skepticism about models [of COVID-19], and they are only as good as the assumptionsyou put into them, but they are not completely misleading. They are telling you something that is a reality, that when you have mitigation that is containing something, and unless it is down, in the right direction, and you pull back prematurely, you are going to get a rebound of cases*.

## 2 Formulation of problem

We consider a standard SIR model for epidemics, which consists of three groups of individuals: those who are *susceptible* and can be passed on the pathogen by the *infectious* individuals, and the *removed* individuals, who are have either developed immunity after infection or who have died. In SIR models, one does not include a flow back from individuals into the susceptible compartment. On longer time-scales, one may also allow for the fact those individuals in the removed group who are immune may eventually return to the susceptible population, which would happen if immunity is only temporary or if a pathogen has evolved substantially. The numbers of individuals in the three classes will be denoted by *S, I*, and *R* respectively, and hence the name “SIR” model. See Figure 1, where we use the symbol “⊗” to indicate that the number of new infected will depend both on the number of susceptibles and infectious (specifically, it will be a product in the classical SIR model to be discussed next, with a proportionality constant *β*), and *ν* denotes the flow to the recovered compartment. Observe that the “feedback” term implicit in the ⊗ effect means that this is not exactly a compartmental system, because for those, *the flow into* a compartment does not depend on the number of individuals in that compartment.

**Figure 1:**
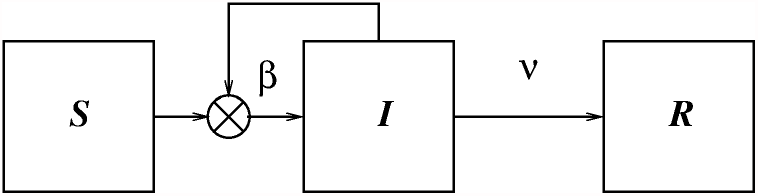
Diagram of standard SIR system

**Figure 2:**
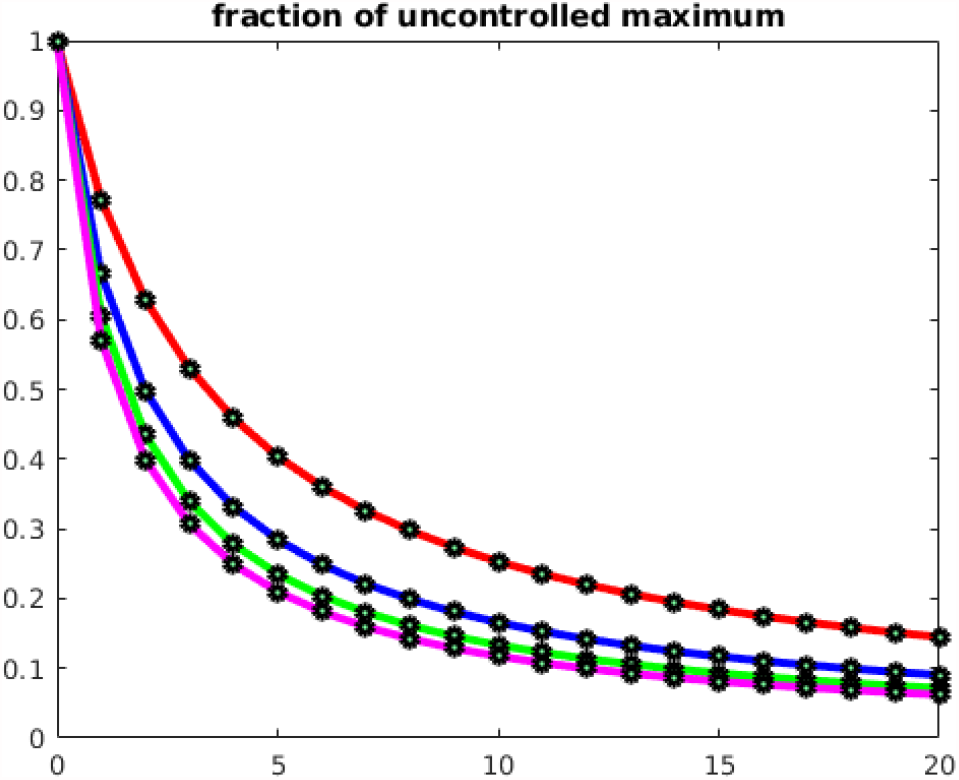
Percentage of virtual peak when using *K* lockdowns (horizontal axis) each of them 7 (red), 14 (blue), 21 (green), or 28 (magenta) days long.

**Figure 3:**
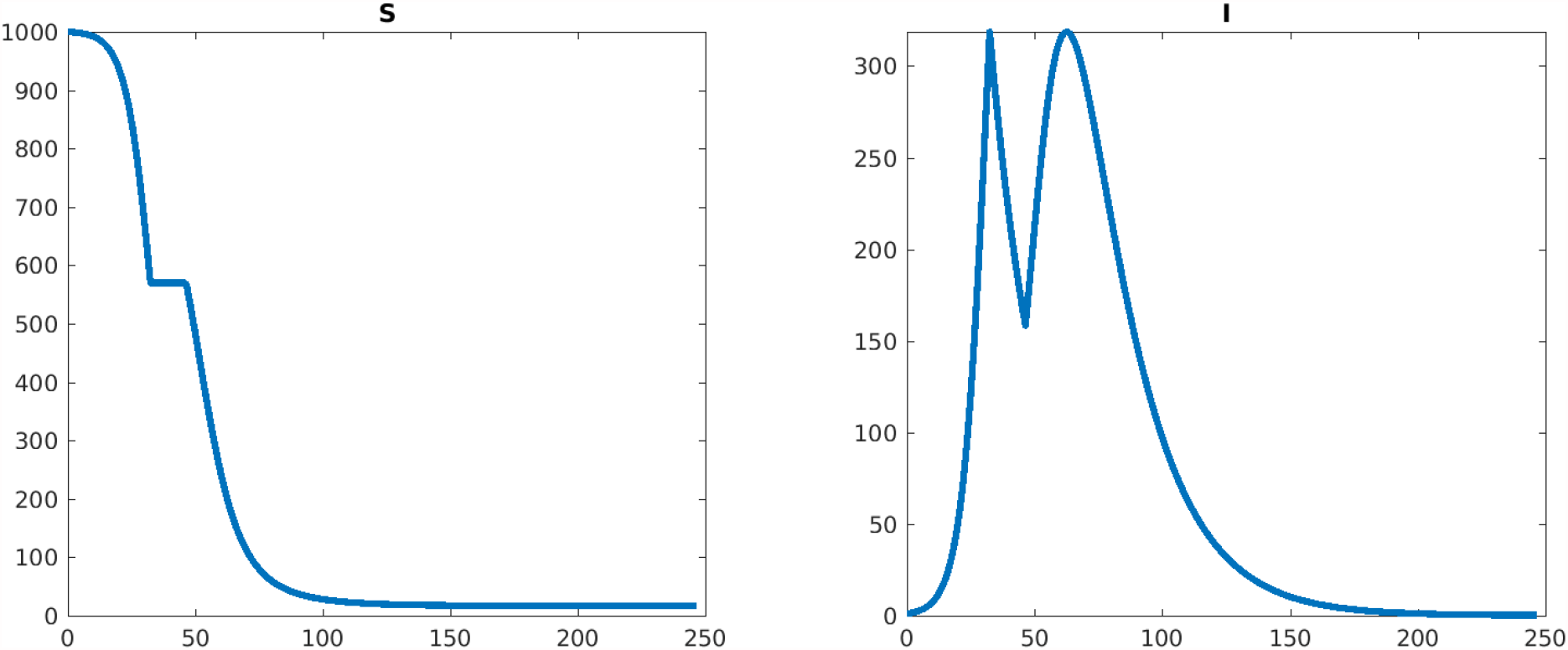
Plots of *S*(*t*) and *I*(*t*) (horizontal axis is time). Number of lockdowns: 1, peak *I* from formula: 318.682808, maximum of *I* on last (no lockdown) period: 318.601530. Lockdown start time(s): 32.42.

**Figure 4:**
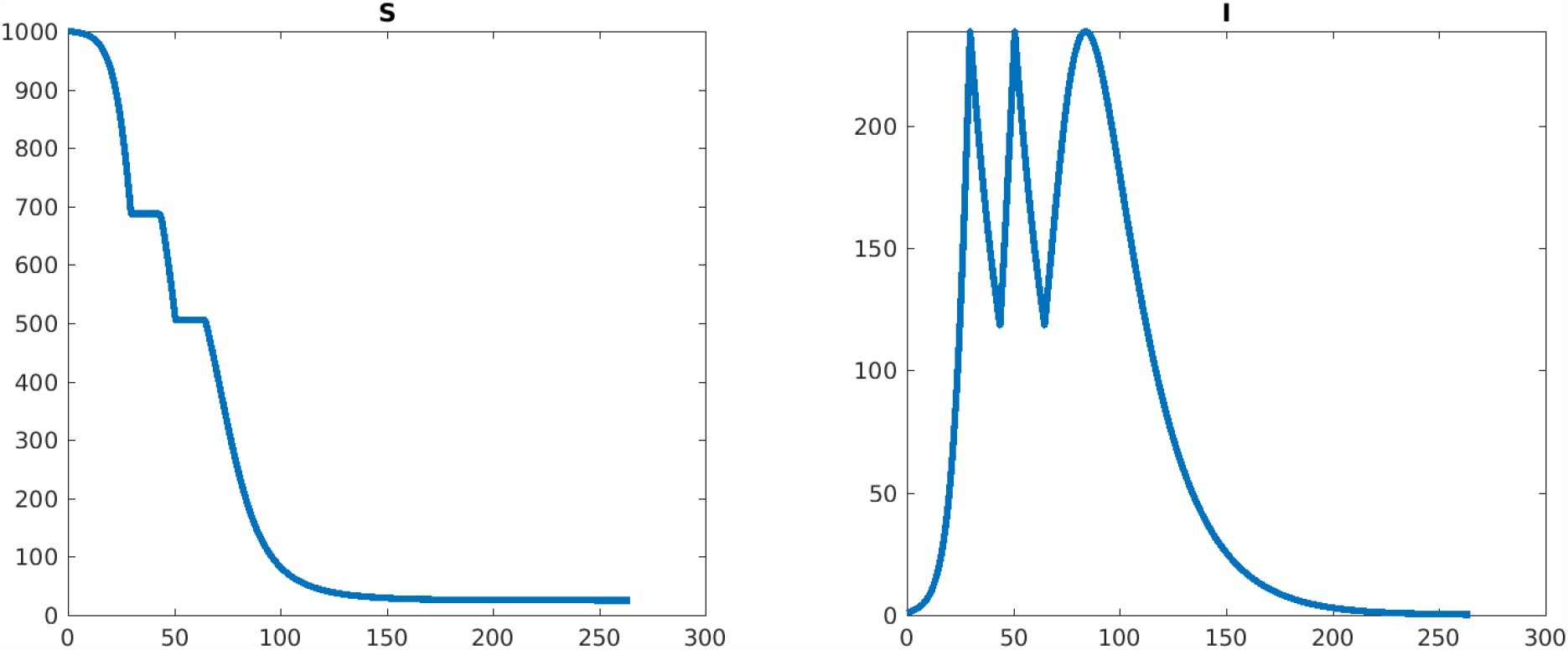
Plots of *S*(*t*) and *I*(*t*) (horizontal axis is time). Number of lockdowns: 2, peak *I* from formula: 238.740981, maximum of *I* on last (no lockdown) period: 238.755779. Lockdown start time(s): 29.73, 50.69.

**Figure 5:**
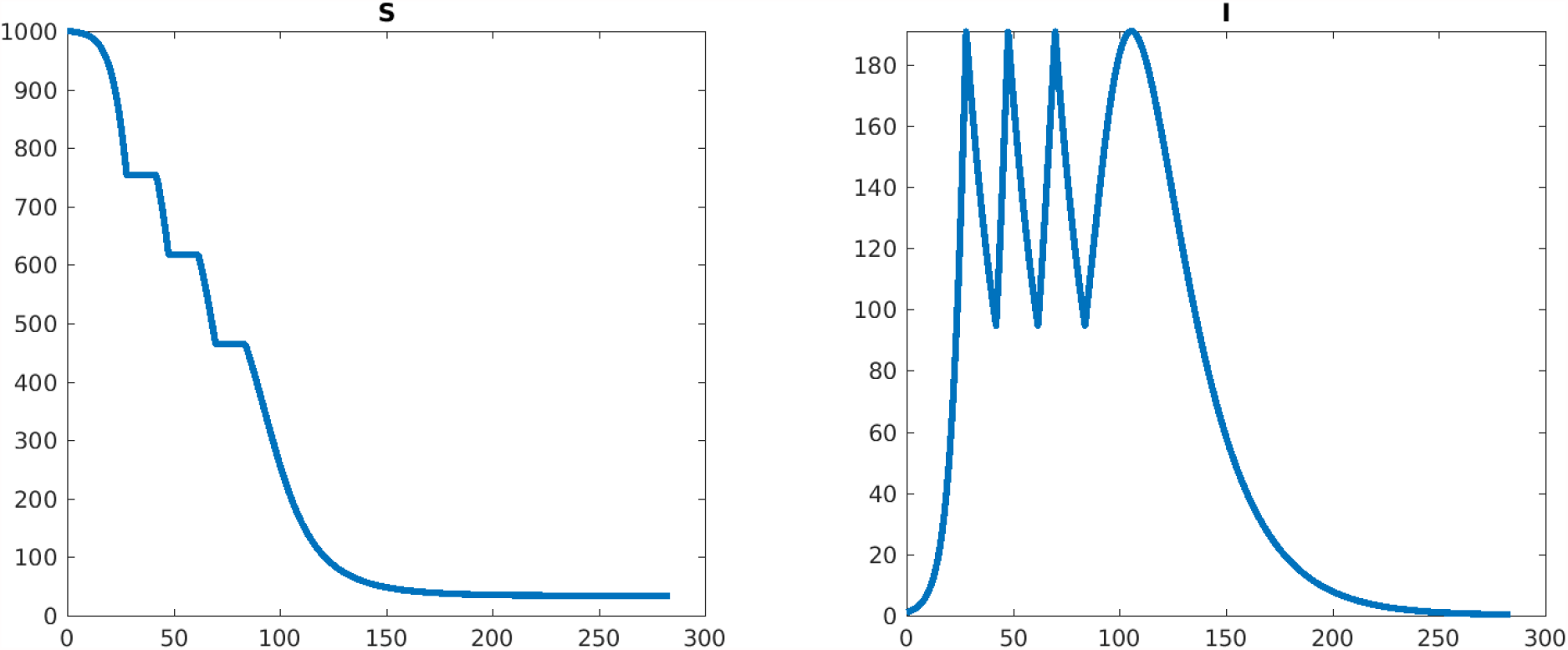
Plots of *S*(*t*) and *I*(*t*) (horizontal axis is time). Number of lockdowns: 3, peak *I* from formula: 190.862880, maximum of *I* on last (no lockdown) period: 191.019085. Lockdown start time(s): 28.01, 47.71, 69.80.

**Figure 6:**
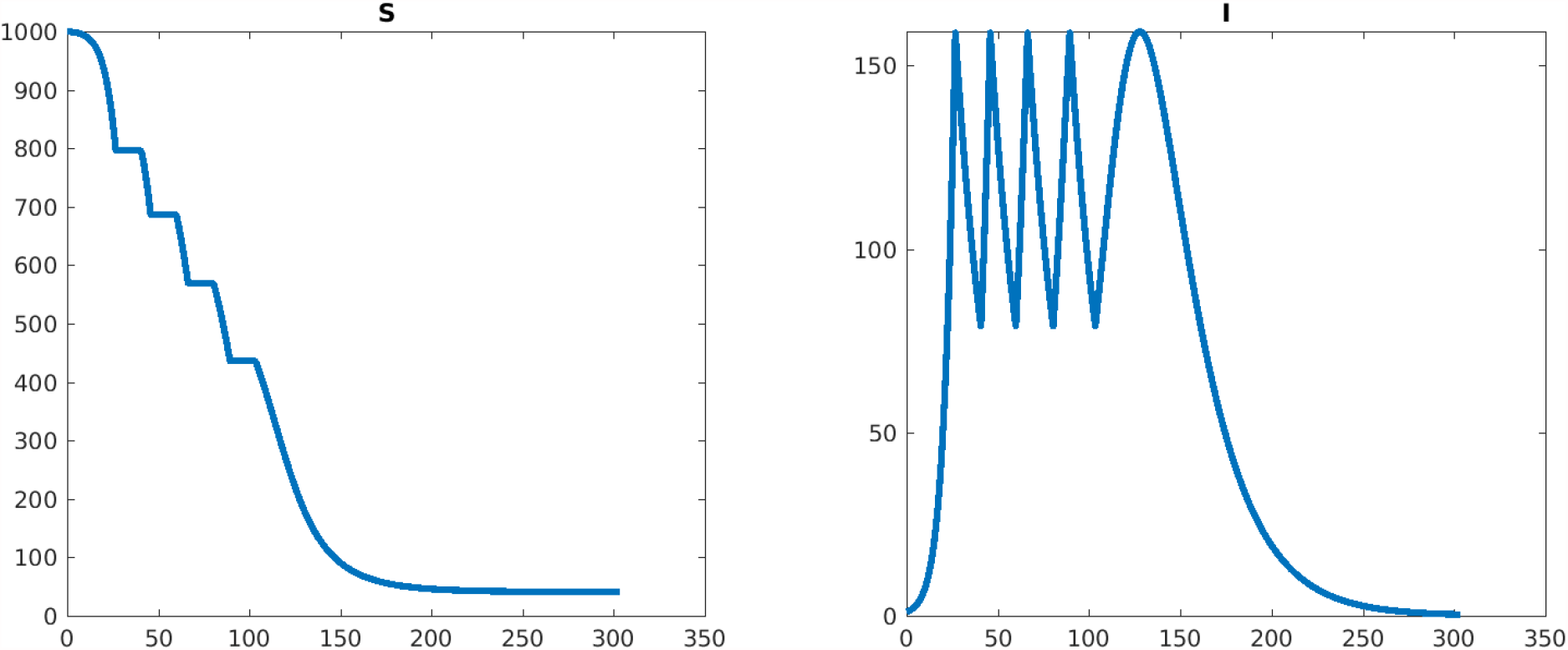
Plots of *S*(*t*) and *I*(*t*) (horizontal axis is time). Number of lockdowns: 4, peak *I* from formula: 158.980313, maximum of *I* on last (no lockdown) period: 159.342577. Lockdown start time(s): 26.74, 45.87, 66.33, 89.44.

**Figure 7:**
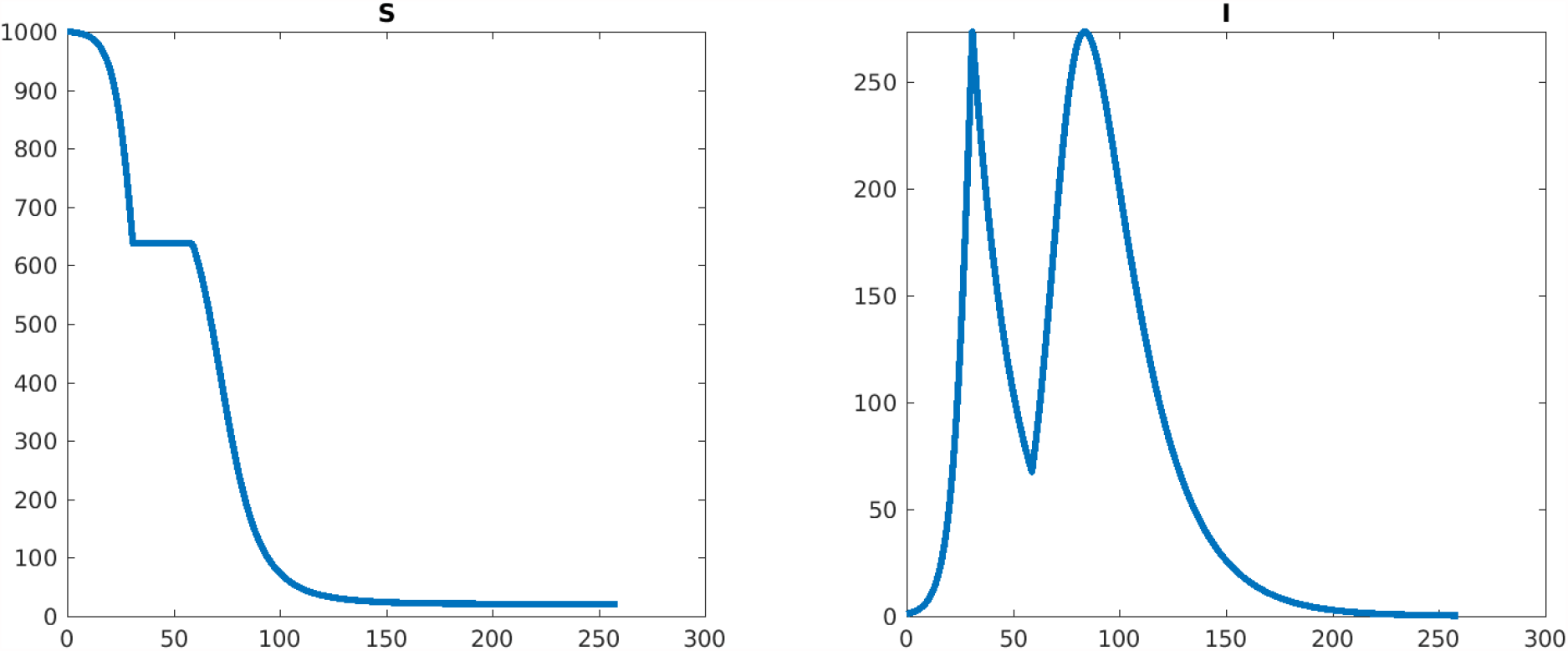
Plots of *S*(*t*) and *I*(*t*) (horizontal axis is time). Number of lockdowns: 1, peak *I* from formula: 273.247170, maximum of *I* on last (no lockdown) period: 273.286548. Lockdown start time(s): 30.9.

**Figure 8:**
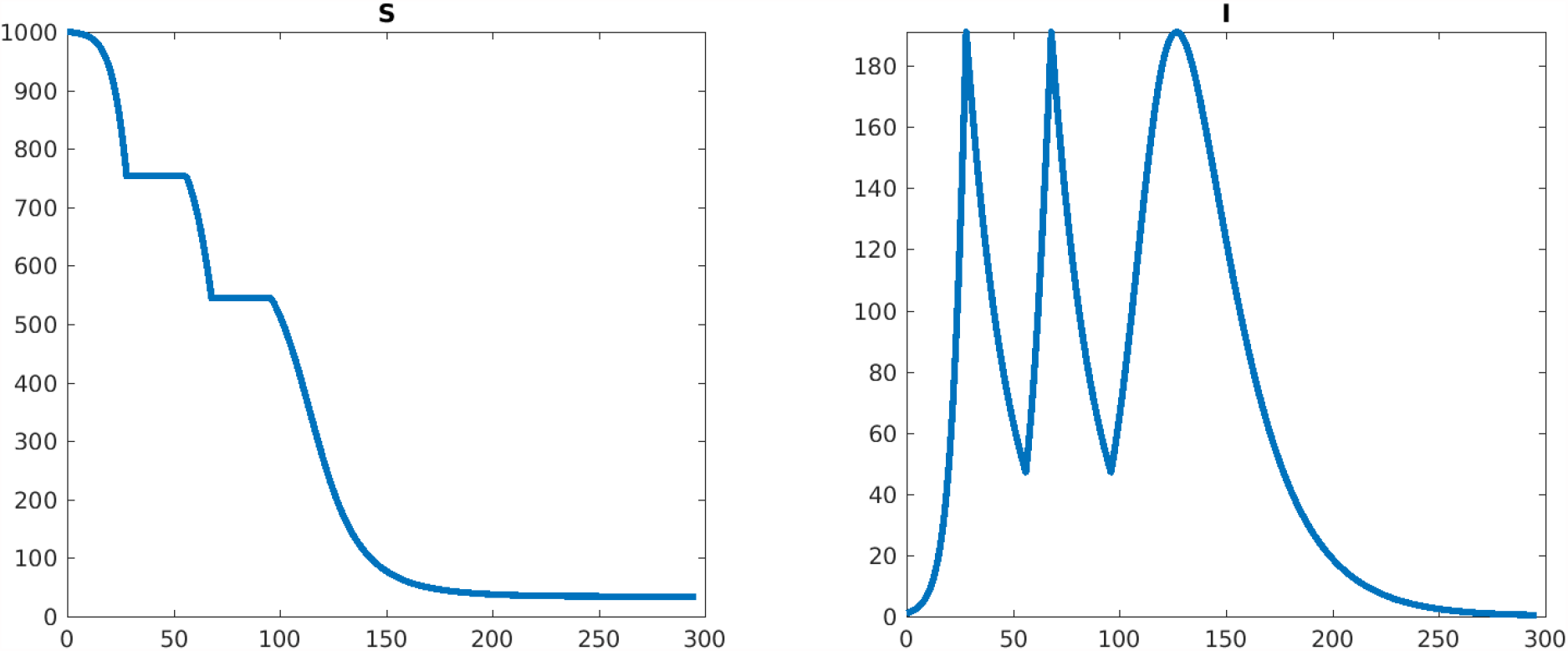
Plots of *S*(*t*) and *I*(*t*) (horizontal axis is time). Number of lockdowns: 2, peak *I* from formula: 191.124644, maximum of *I* on last (no lockdown) period: 191.048654. Lockdown start time(s): 28.02, 68.02.

**Figure 9:**
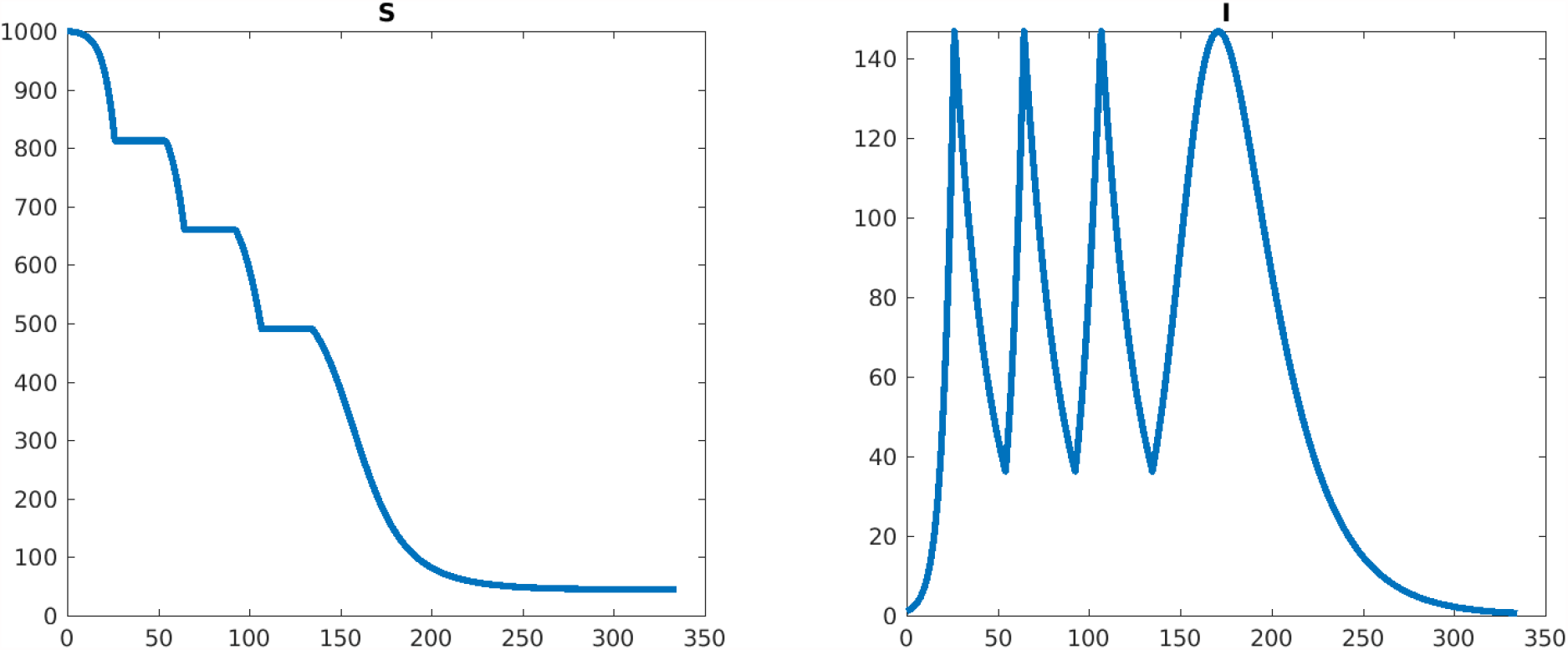
Plots of *S*(*t*) and *I*(*t*) (horizontal axis is time). Number of lockdowns: 3, peak *I* from formula: 146.957573, maximum of *I* on last (no lockdown) period: 146.913773. Lockdown start time(s): 26.22, 64.39, 106.74.

**Figure 10:**
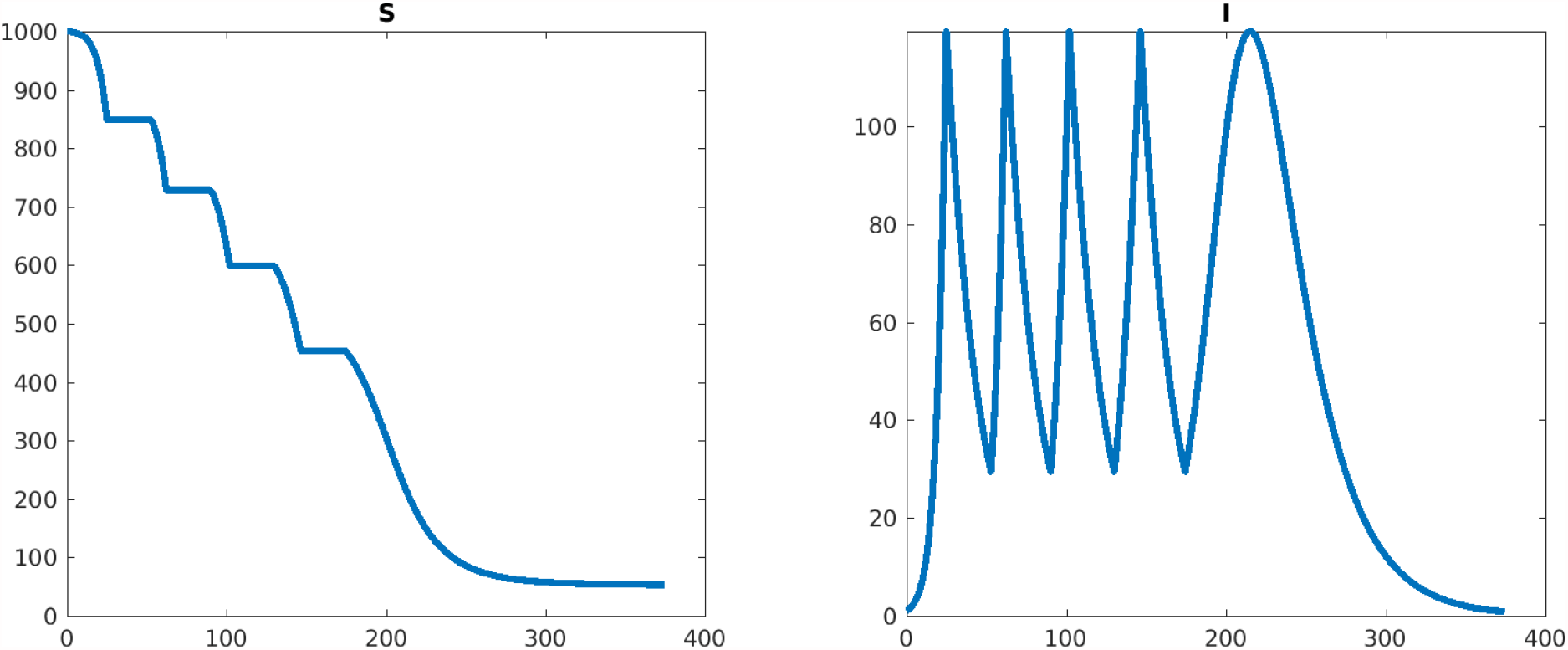
Plots of *S*(*t*) and *I*(*t*) (horizontal axis is time). Number of lockdowns: 4, peak *I* from formula: 119.371878, maximum of *I* on last (no lockdown) period: 119.395364. Lockdown start time(s): 24.91, 62.23, 101.98, 146.47.

Mathematically, assuming that new infections are due to contacts between *S* and *I* individuals, and that the rate at which this happens is proportional to the numbers of such individuals, there results a system of three coupled ordinary differential equations as follows:

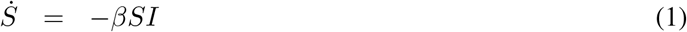

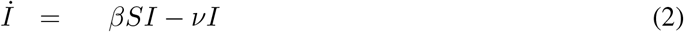

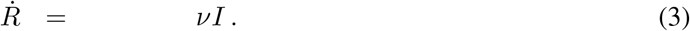

From now on we will ignore the last equation since it does not affect the number of infectives. The initial state satisfies *S*(0) = *S*_0_ > 0, *I*(0) = *I*_0_ > 0. The positive parameter *β* (infectivity/contact rate) quantifies the transmission rate between the susceptible *S* and infected *I* individuals in a well-mixed population, and *ν* (recovery/death rate) is the rate of flow into the removed *R* compartment. We will assume that ℛ_0_ := *βS*_0_*/ν* > 1; otherwise, the problem to be discussed is not interesting, as *I* decreases monotonically to zero if the condition does not hold. (This and other well known facts about SIR systems are reviewed in Section 5; see also e.g. [19].)

In the SIR model, NPIs are viewed as reducing the contact rate *β*. (More sophisticated models of social distancing have been widely studied, see for example [20, 21, 22, 23, 24, 25, 26].) The reduction of *β* is modeled by a time-varying *β*(*t*) where

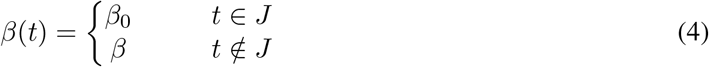

where 0 ≤ *β*_0_ < *β* are two fixed values, and *J* ⊆ [0, ∞). In our work, *J* will be the union of a number *K* of intervals

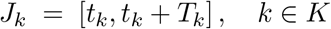

and *we will be taking β*_0_ = 0, representing a strict or full lockdown. The lengths *T*_*k*_ of the respective intervals will be allowed to be arbitrary (but fixed) in our theory, though the most elegant formulas are obtained when they are all equal (we might have *T*_*k*_ = 14 or 28 days for all *k*, for instance). The objective will be to minimize the maximum of *I*(*t*) for *t* ≥0 (“flatten the curve”) by appropriately choosing the start times *t*_*k*_.

As discused in the introduction, even if perfect lockdowns are not completely realistic, studying this case helps understand the case *β*_0_ > 0. Indeed, we show via simulations that the conclusions for *β*_0_ = 0 are very relevant to the case of a small but nonzero *β*_0_, even one that is about 20% of the value of *β*.

Previous work on this and related problems includes [27], which treats an optimal schedule minimizing a combination of the total number of deaths and the peak of the infected compartment, [17] which shows that a single interval (*K* = 1) is optimal if the objective is to minimize the total number of susceptible individuals at the end of the epidemic, [15] with numerical studies of optimally timing fixed-duration “one-shot” strategies, and the very nice theoretical paper [16] which showed that the optimal strategy for minimizing peak infection is a combination of a strict lockdown (“full suppression”) with a feedback strategy which keeps ℛ_0_=1. Also closely related to this work is [28], which studies timing of lockdowns, including periodic strategies, through a combination of theoretical and numerical methods. Periodic strategies are also studied in [25, 29, 30, 31] as well as other references.

### 2.1 Main result

Let

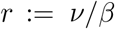

and consider the “virtual peak” of *I*(*t*) if no lockdown were imposed:

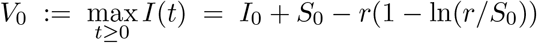

(cf. Section 5). Define this expression:

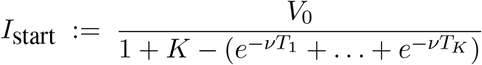

The main result is as follows. It considers the optimization problem in which the number of lockdowns is fixed (*K*) as well as their respective lengths (*T*_*k*_’s). The parameters to be optimized over are the times at which the each lockdown should commence.

#### Theorem 1

Suppose that *I*(0) < *I*_start_. Then, in order to minimize the maximum of *I*(*t*), *t* ≥0, lockdowns should start whenever

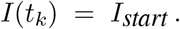

Moreover, under this policy, the maximum *I*_max_ of *I*(*t*) will equal *I*_*start*_.

In other words, any time that the infective population level reaches the value *I*_start_, the next time-*T*_*k*_ lockdown interval *I*_*k*_ should start.

For example, if *T*_*k*_ = *T* for *i* = 1, …, *K* then the formula is

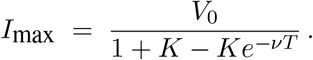

Note that as *T* → ∞ the best possible maximum peak is

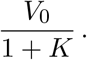

**Proof of Theorem 1:** For mathematical elegance, we will include the theoretical possibility of a lockdown starting at time exactly *t*_0_ = 0 (and later make *T*_0_ = 0, so that there is in effect no initial lockdown).

For any initial population (*σ, ι*), we introduce the following function:

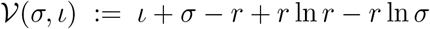

which gives the peak value of *I*(*t*) if we start at the initial population (*σ, ι*) and there would be no further lockdowns, and let:

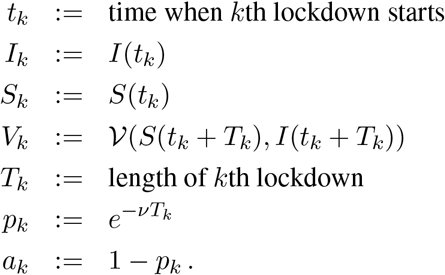

(so *t*_0_ = 0 and (*S*_0_, *I*_0_) is the state at the start of the epidemic). Observe that, for all *k*,

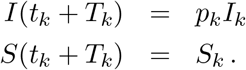

Consider these equalities:

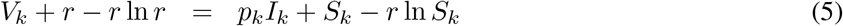

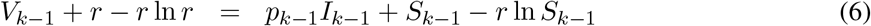

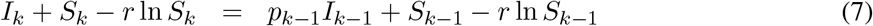

where the last equality follows from the conservation law *I*(*t*) + *S*(*t*) −*r* ln *S*(*t*) ≡constant, applied to a solution that starts at *ι* = *p*_*k*_*I*_*k*_ and *σ* = *S*_*k*_. Adding (5) to (7) and subtracting (6), we obtain:

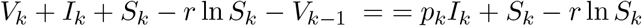

from which we conclude the following recursion:

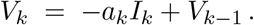

Applying this formula recursively, we conclude:

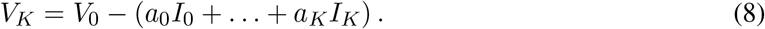

The largest value of *I*(*t*) will occur at the maximum of the peaks occurring at the start of each *k*th lockdown, or the last “virtual peak” (which is then a real local maximum). In other words, the maximum of *I* is

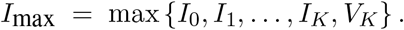

Under the epidemic assumption ℛ_0_ > 1, *İ*(0) > 0, so we can drop *I*_0_ from the list. We replace the value for *V*_*K*_ from formula (8). Letting *T*_0_ = 0 means that *a*_0_ = 1 − *p*_0_ = 0, so we conclude that

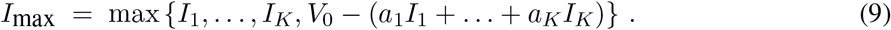

To minimize the peak, we need to pick non-negative *I*_1_, …, *I*_*K*_ in such a way that this expression is minimized. This is achieved at a unique global minimum, at which

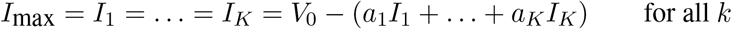

(see Lemma 1 below), namely

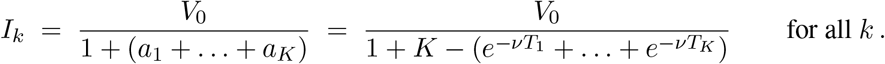

This completes the proof of Theorem 1. ▪

#### Lemma 1

*For any a*_1_ ≥ 0, …, *a*_*K*_ ≥ 0 *and b* > 0, *define*

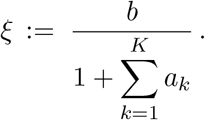

*Consider the following function, defined on* 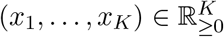:

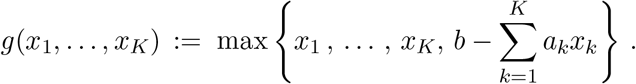

*Then, the minimum value of g is ξ, and it is achieved at the unique point where all x*_*k*_ = *ξ*.

**Proof of Lemma 1:** Define 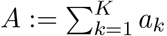, so that 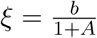, and note these properties:

1. *g*(*ξ*, …, *ξ*) = *ξ*. To prove this, note that, when all *x*_*k*_ = *ξ*,

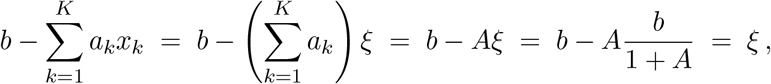

so all terms in the max are the same.
2. If *g*(*x*_1_, …, *x*_*K*_) ≤ *ξ* then *x*_*k*_ = *ξ* for all *k*. Notice first that *x*_*k*_ ≤ max {*x*_1_, …, *x*_*K*_} ≤ *g*(*x*_1_, …, *x*_*K*_) ≤ *ξ* for all *k*. If for some *k* this inequality were strict, then −*x*_*k*_ ≥ −*ξ* for all *k*, and −*x*_*k*_ > −*ξ* for some *k*, so

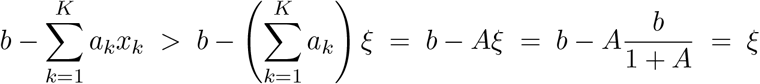

but and this would contradict 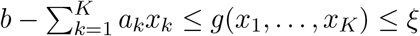.

Property 2 shows that *g*(*x*_1_, …, *x*_*K*_) > *ξ* unless all *x*_*k*_ = *ξ*, and Property 1 shows that the minimum is achieved when all *x*_*k*_ = *ξ*. ▪

We next show some illustrations of the use of the formula for several lengths of lockdowns as well as simulations for various scenarios of 1, 2, 3, or 4 lockdowns. We use reasonable parameters in each case. Finally, we will show computationally that a small positive *β*_0_ does not change conclusions much, at least for the case of a single lockdown.

### 2.2 Equal lengths are optimal

The formula derived in this paper allows one to prove the following result. Suppose that we are given a total “lockdown time budget” *T* > 0, and wish to find *K* lockdown times *T*_*k*_, such that 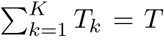 so as to minimize 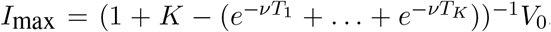. Minimizing *I*_max_ over the *T*_*k*_’s is equivalent to minimizing

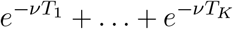

subject to

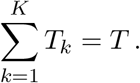

The functions 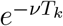 are strictly convex, the objective function is separable, and the constraint is affine, so it follows that there is a unique solution to this optimization problem (see Example 5.4 in [32]). Consider the Lagrangian:

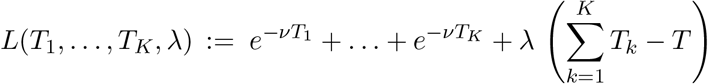

and set the *K* partial derivatives with respect to the *T*_*k*_’s to zero. This gives:

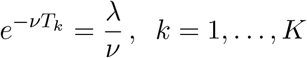

which implies *T*_1_ = … = *T*_*K*_. From 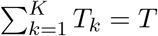 it follows that

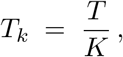

i.e. equal lengths are optimal.

We now prove that *the more intervals, the better*. Indeed, suppose that we compare the optimal solution with *K* intervals to the optimal solution with *K*+1 intervals. Let us think of the optimal solution with *K* intervals as a particular solution with *K*+1 intervals, in which the last interval is zero (and the first *K* are equal to *T/K*). Clearly, this last solution is not optimal, since the (unique) optimal solution to the problem with *K*+1 intervals has all intervals nonzero (and equal to *T/*(*K*+1)). In other words, *having one more interval is always better*. Of course, this ignores the social, economic, and psychological problems of imposing multiple lockdowns.

A variation of the “total budget” problem is as follows. Suppose that the “cost” of a second lockdown is different from (and presumably larger than) the cost of the first lockdown. This may represent “lockdown fatigue” or lack of political will. More generally, each subsequent lockdown could have an associated cost *c*_*k*_ > 0. In this case, the interesting mathematical problem would be to again minimize 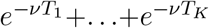 but this time subject to a general affine constraint

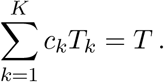

The Lagrangian is now

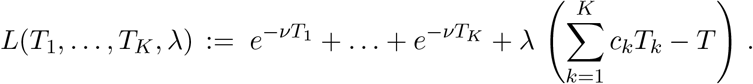

We normalize the problem by asking 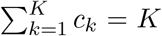, so that the equal-cost case is that when all *c*_*k*_ = 1. Proceeding as before, we arrive at the equations:

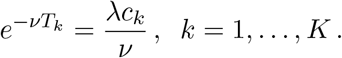

Pick any index *k* ≠ 1. Then

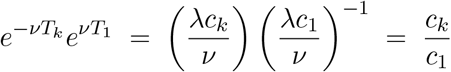

or 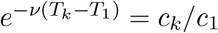 from which we derive the formulas

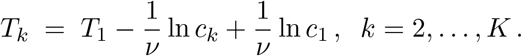

Thus:

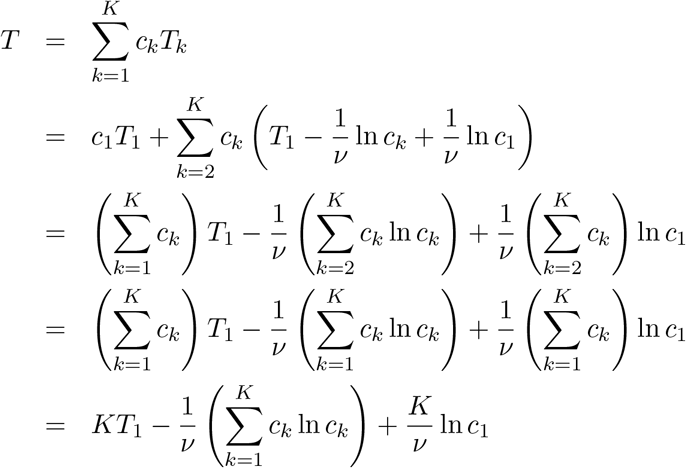

(using that 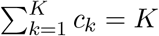). We conclude:

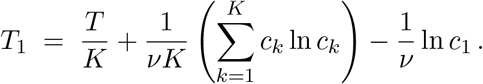

It follows that, for each *i* ≥ 2, we have an analogous formula:

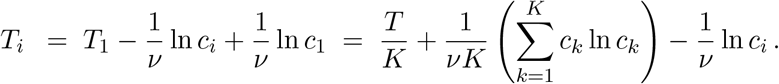

Note the common entropy-like term as well as the last term that makes intervals shorter when their “cost” is higher. In the special case that all *c*_*k*_ = 1, the logarithms are zero and we recover the previous case *T*_*i*_ = *T/K*.

### 2.3 Sensitivity to parameters

The recovery rate *ν* is relatively easy to estimate, wince it is largely a function of the physiology of the pathogen. However, the parameter *β* depends on behavioral characteristics, population density, and so forth, so it is much harder to know accurately. A natural question to ask if: how much worse does a policy based on an incorrect value perform, relative to the optimal policy that would have been used if *β* were perfectly known? We study this sensitivity question here, restricting for simplicity to the case of a single (perfect) lockdown, i.e. *K* = 1, *T*_1_ = *T*.

Suppose that the population value *I*_1_ at which the lockdown starts was optimized based on a “wrong” value 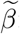:

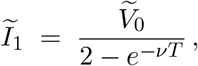

where

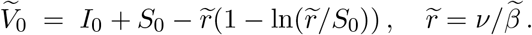

The maximum value of *I*(*t*) is:

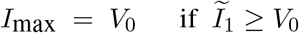

(because the maximum with no lockdowns is *V*_0_, so the lockdown will never be triggered in the case *Ĩ*_1_ ≥ *V*_0_) and

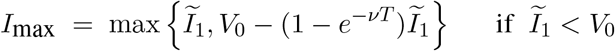

(formula 9), where

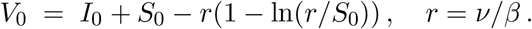

In contrast, the optimal solution would have been to pick

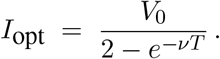

Since this is optimal, of course *I*_max_ ≥*I*_opt_. Let us consider the relative penalty (in terms of maximum infectives) incurred by using the wrong *β*:

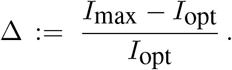

Let is write *p* := *e*^−*νT*^ to simplify notations. When *Ĩ*_1_ ≥ *V*_0_ (that is, if 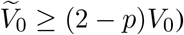):

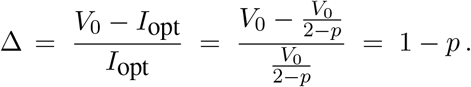

When *Ĩ*_1_ < *V*_0_ (that is, if 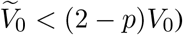):

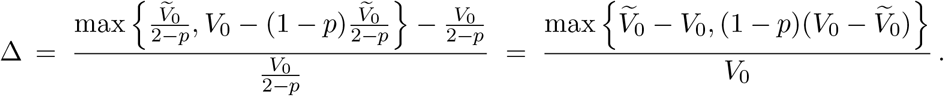

In this maximum, the terms have opposite sign, so the max is achieved at the non-negative one.

A formula for Δ can be most conveniently derived when thinking of Δ as a function of 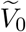 (with *β*, and therefore also *V*_0_, fixed). We conclude:

#### Theorem 2

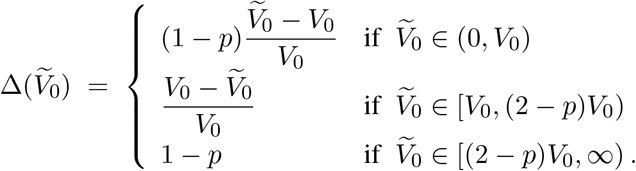

From this formula, one can immediately compute sensitivity to 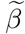. Observe that 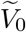 is a strictly increasing function of 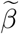, because the derivative of the function 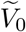 with respect to 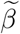is:

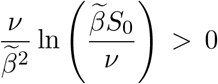

as long as we are in the epidemic parameter range ℛ_0_ := *βS*_0_*/ν* > 1.

Note that Δ is continuous on 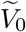; it is zero (no penalty) when 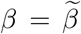, i.e. 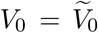, and 1 − *p* when substituting 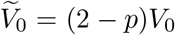 into 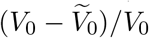. In the limit that *p* → 1 (that is, if the lockdown interval *T* → 0, the middle case becomes vacuous and the sensitivity is zero (as expected).

## 3 Some numerical explorations

### 3.1 Optimal reduction of “virtual peak” with *K* perfect lockdowns

The use of a even a small number of lockdowns results in a drastic reduction of the peak that would occur (the “virtual peak”) if there were no lockdowns. The marginal benefit of additional lockdowns is relatively minor, after a certain number of them, as is the benefit of longer lockdowns. We plot here the fraction (1 + *K*(1 − *e*^−*νT*^))^−1^ for *ν* = 0.05 and *T* = 7, 14, 21, 28 days.

## 4 Simulations

### 4.1 Simulations using optimal formula

We simulate various lockdown lengths using our optimal formulas. Parameters are, *β* = 0.00025, *ν* = 0.05 (so ℛ_0_ = 5), initial conditions are *S*_0_ = 1000, *I*_0_ = 1, and the peak if there are no lockdowns is *I* = 479. The maximum from the formula coincides with the maximum in simulations, up to numerical error.

#### 4.1.1 Lockdown length is *T* = 14 days, perfect lockdown

#### 4.1.2 Lockdown length is *T* = 28 days, perfect lockdown

### 4.2 Comparing multiple vs. single lockdowns

We proved that more lockdowns are always better, if the total lockdown “budget” is fixed. Here we show the following comparisons: one 28-day compared to two 14-day lockdowns, and two 28-day compared to four 14-day lockdowns (Figures 11 and 12 respectively). Observe that the peaks when using repeated lockdowns are lower, as theory predicts. Interestingly, the timings of the last peak and asymptotic behaviors look identical in the respective left and right plots.

**Figure 11:**
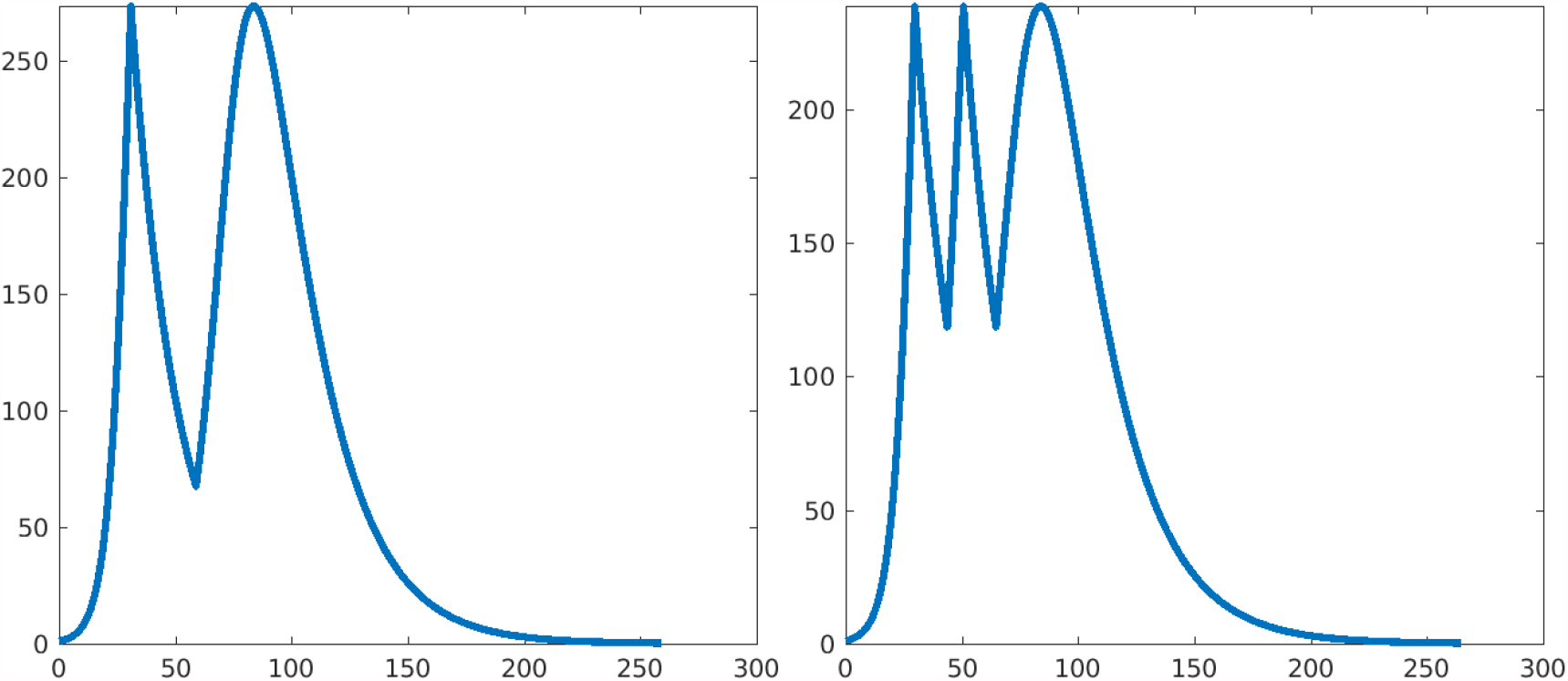
Left: one 28-day lockdown. Right: two 14-day lockdowns. Parameters as earlier.

**Figure 12:**
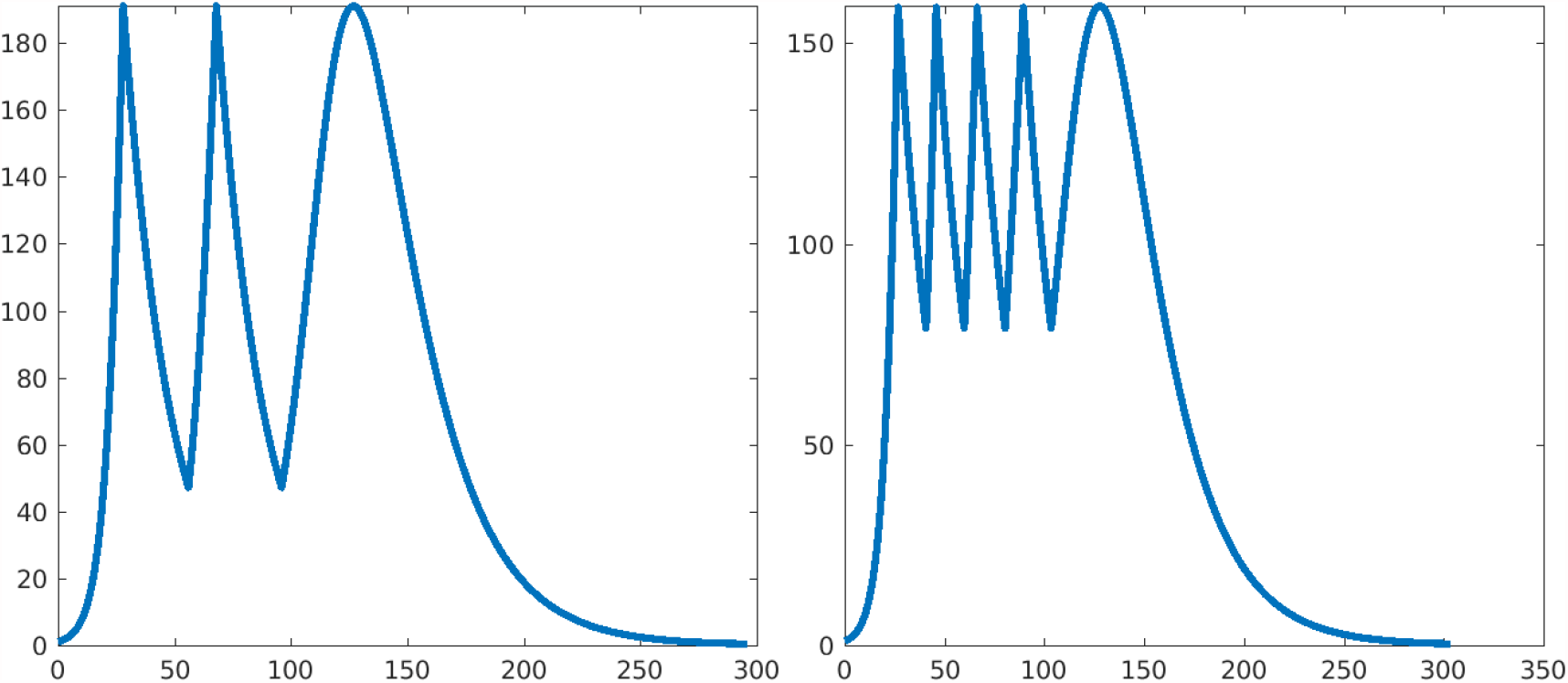
Left: two 28-day lockdowns. Right: four 14-day lockdowns. Parameters as earlier.

**Figure 13:**
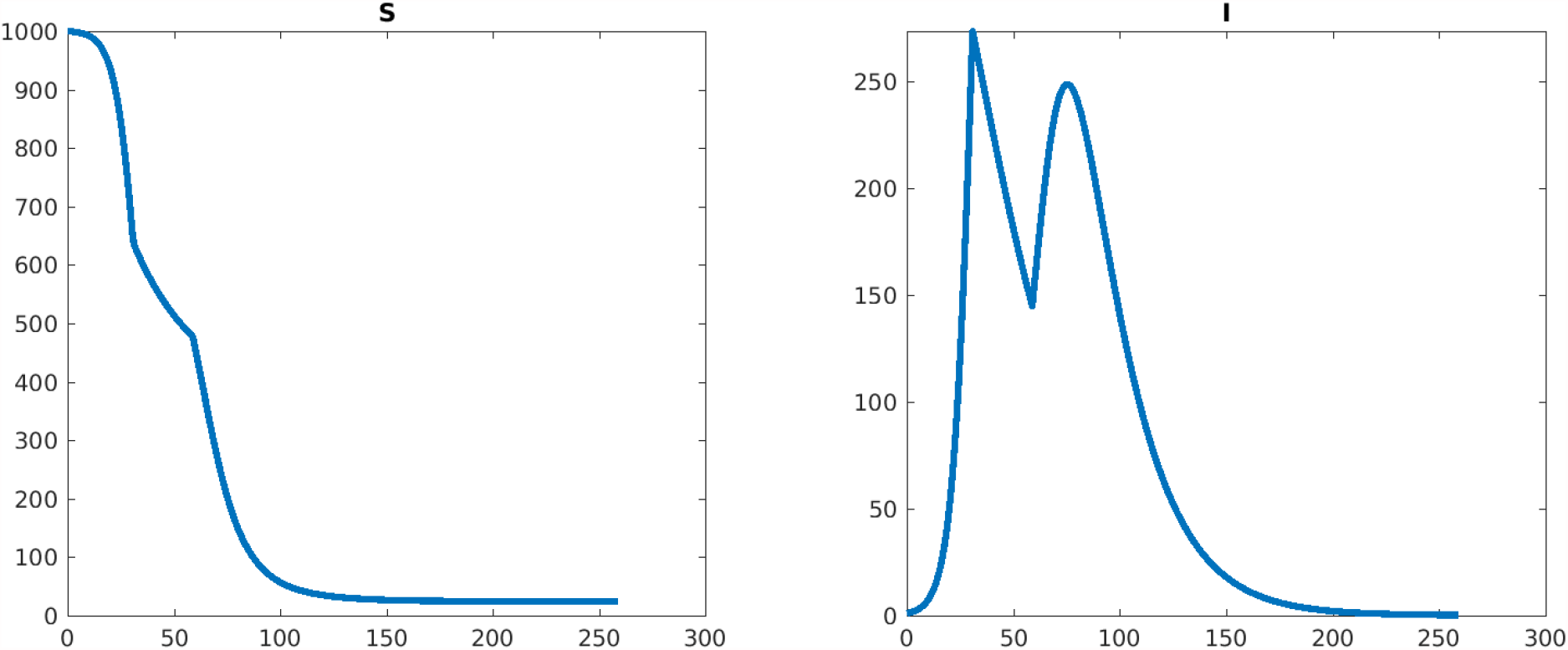
Plots of *S*(*t*) and *I*(*t*) (horizontal axis is time). Number of lockdowns: 1, peak *I* from formula: 273.247170, maximum of *I* on last (no lockdown) period: 248.383407. Lockdown start time(s): 30.90.

**Figure 14:**
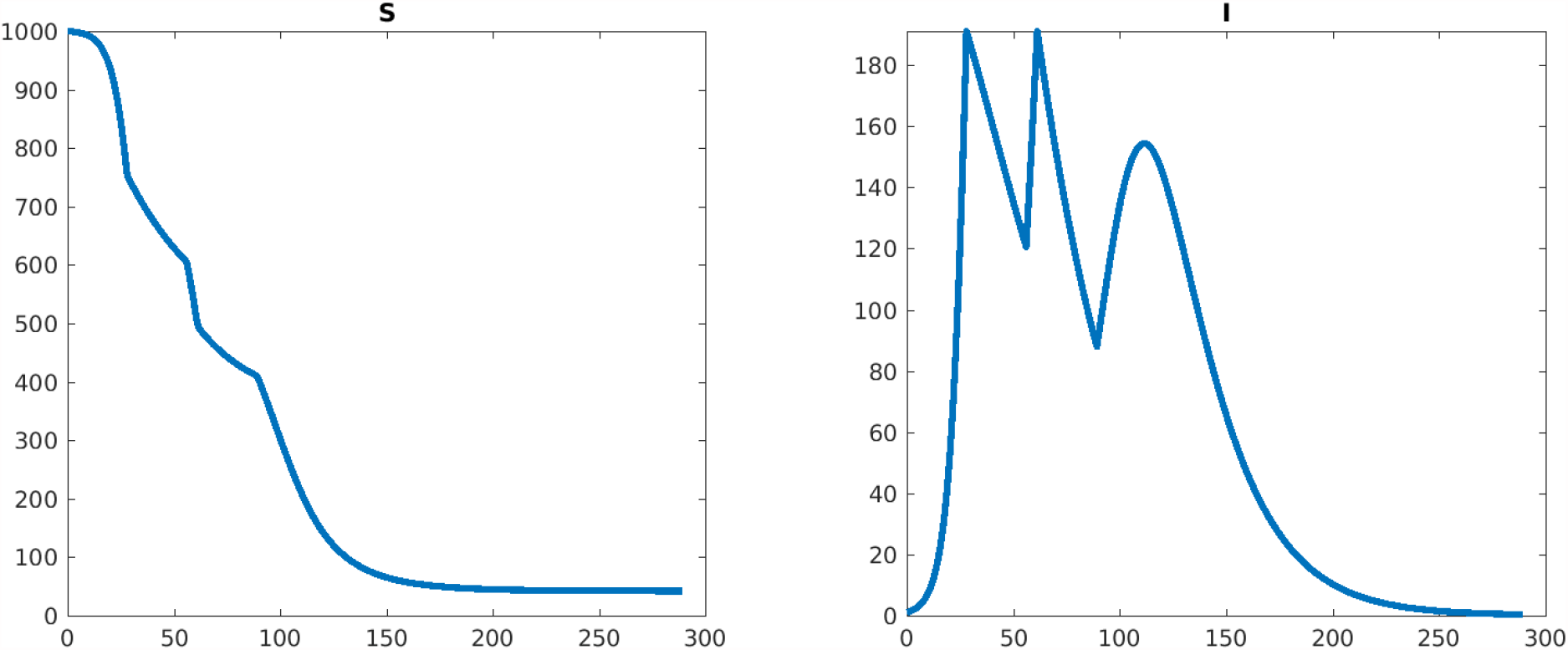
Plots of *S*(*t*) and *I*(*t*) (horizontal axis is time). Number of lockdowns: 2, peak *I* from formula: 191.124644, maximum of *I* on last (no lockdown) period: 154.387915. Lockdown start time(s): 28.02, 61.2.

**Figure 15:**
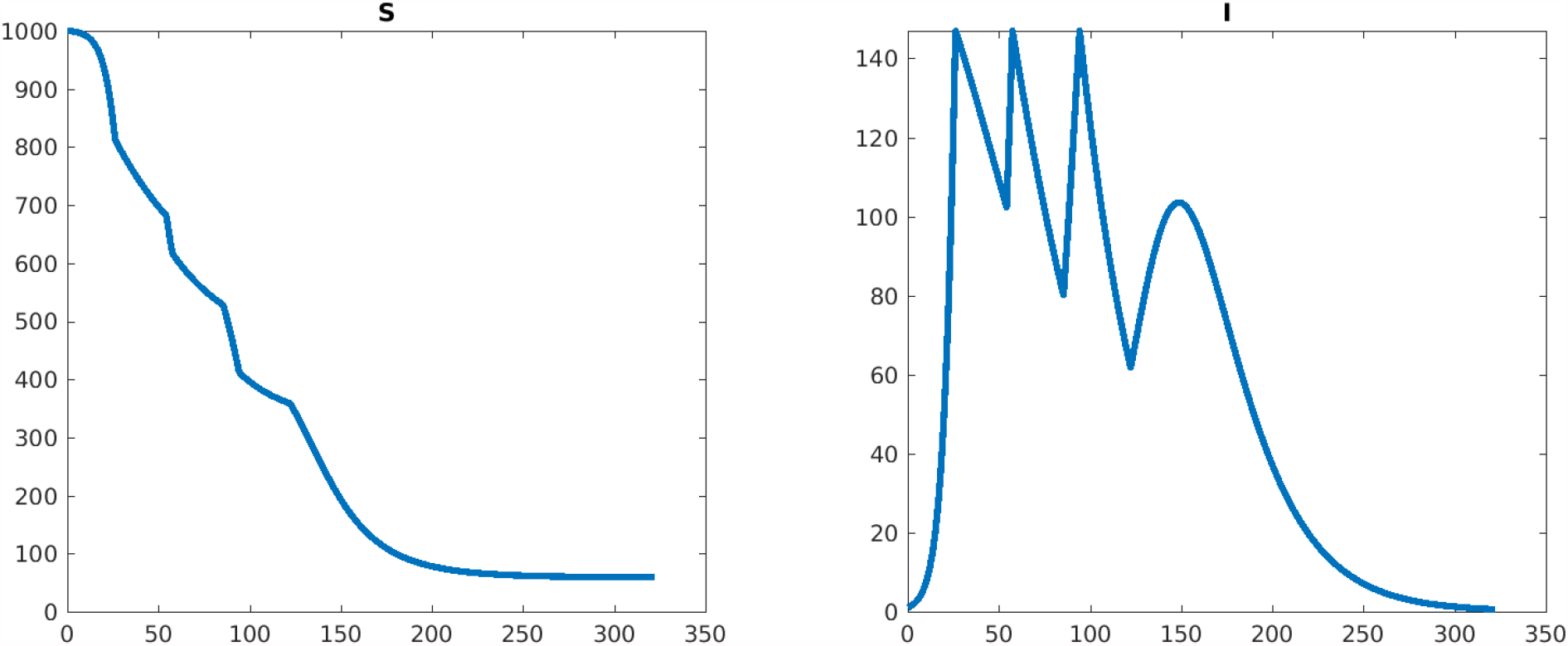
Plots of *S*(*t*) and *I*(*t*) (horizontal axis is time). Number of lockdowns: 3, peak *I* from formula: 146.957573, maximum of *I* on last (no lockdown) period: 103.506053. Lockdown start time(s): 26.22, 57.43, 94.30.

**Figure 16:**
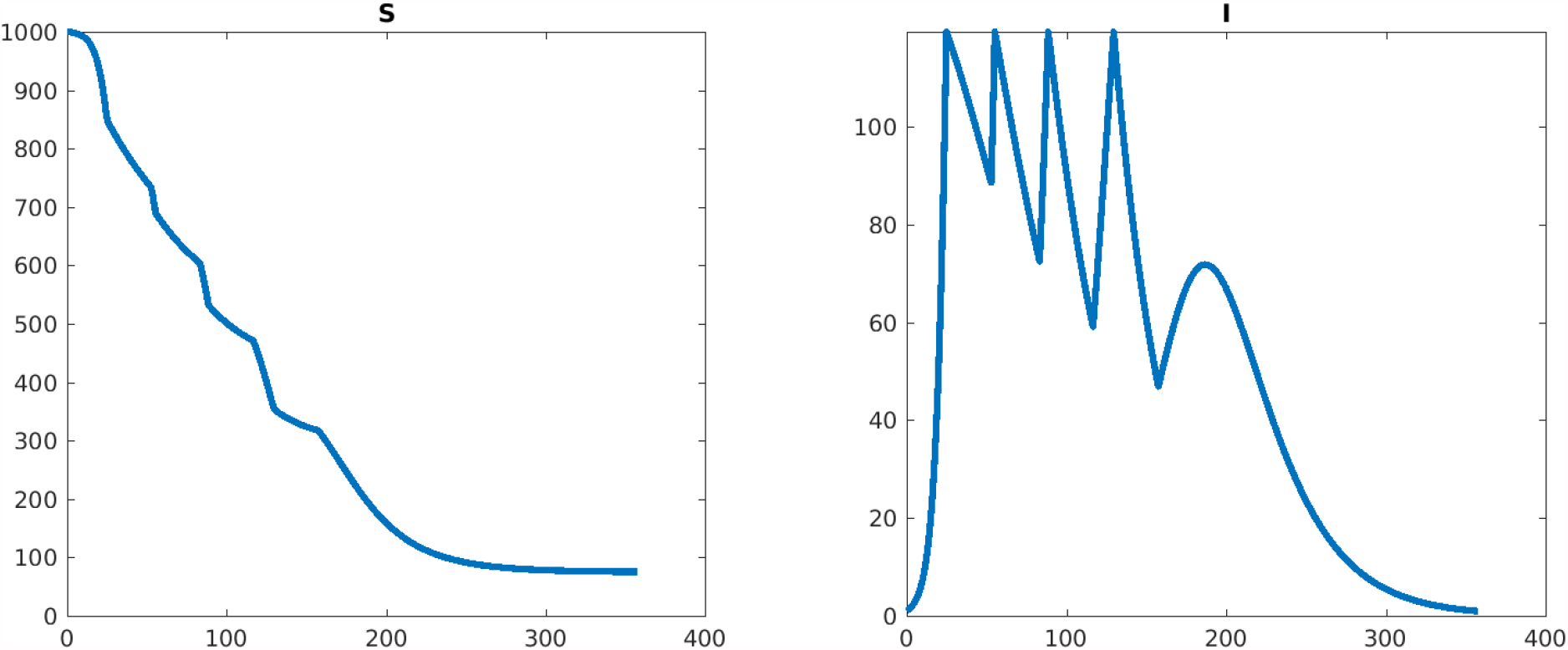
Plots of *S*(*t*) and *I*(*t*) (horizontal axis is time). Number of lockdowns: 4, peak *I* from formula: 119.371878, maximum of *I* on last (no lockdown) period: 71.792718. Lockdown start time(s): 24.91, 55.24, 88.63, 129.63.

### 4.3 Testing *β*_0_ ≠ 0 with formula derived for *β*_0_ = 0

We simulate the use of the formula derived for *β*_0_ = 0, under the lockdown value *β*_0_ = 0.00005, which represents a 20% value of the normal contact rate. Again, *β* = 0.00025, *ν* = 0.05 (so ℛ_0_ = 5), initial conditions are *S*_0_ = 1000, *I*_0_ = 1, and the peak if there are no lockdowns is *I* = 479.

#### 4.3.1 Lockdown length is *T* = 28 days, *β*_0_ = 0.00005

#### 4.3.2 Lockdown length is *T* = 14 days, *β*_0_ = 0.00005

### 4.4 Comparison with optimal strategy for *β*_0_ ≠ 0

For comparison with the use of the optimal formulas derived for *β*_0_ = 0, we show here numerically the optimal solution when *β*_0_≠0, specifically *β*_0_ = 0.00005 as above. As earlier, parameters are *β* = 0.00025, *ν* = 0.05 (so ℛ_0_ = 5), initial conditions are *S*_0_ = 1000, *I*_0_ = 1, and the peak if there are no lockdowns is *I* = 479. We take only the case of a single lockdown, for simplicity, and lockdown lengths of 14 or 28 days.

We find that the formula predicts the optimal timing extremely well for 14-day lockdowns (error less than 1% in maximum infectives), and is fairly good for 28-day lockdowns as well (about 5% error).

#### 4.4.1 Lockdown length is *T* = 14 days, *β*_0_ = 0.00005

For 14-day lockdowns, the plot in Figure 17 suggests that the formula derived for *β*_0_ = 0 triggered the first lockdown too early, so we explored by what fraction > 1 to increase the trigger point, from which the optimal strategy is clear.

**Figure 17:**
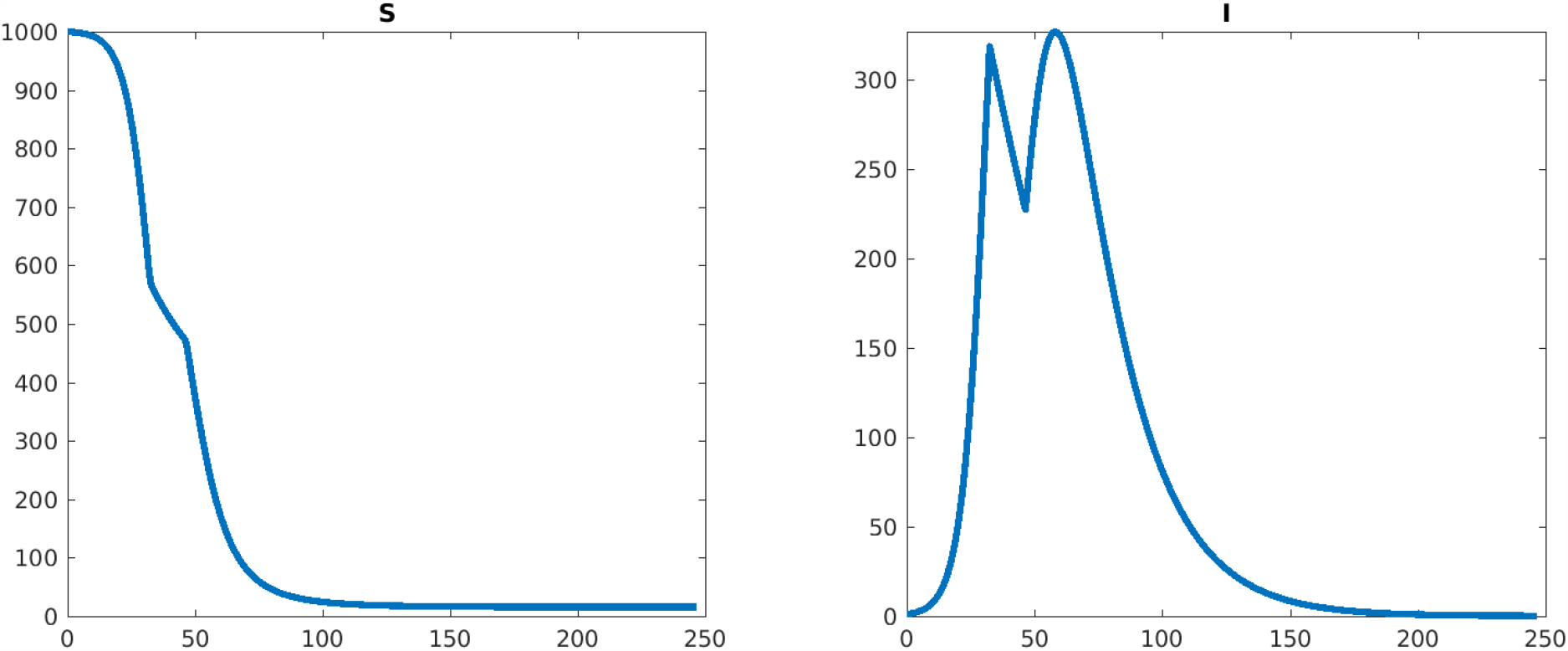
Plots of *S*(*t*) and *I*(*t*) (horizontal axis is time). Number of lockdowns: 1, peak *I* from formula: 318.682808, maximum of *I* on last (no lockdown) period: 326.846639. Lockdown start time(s): 32.4.

**Figure 18:**
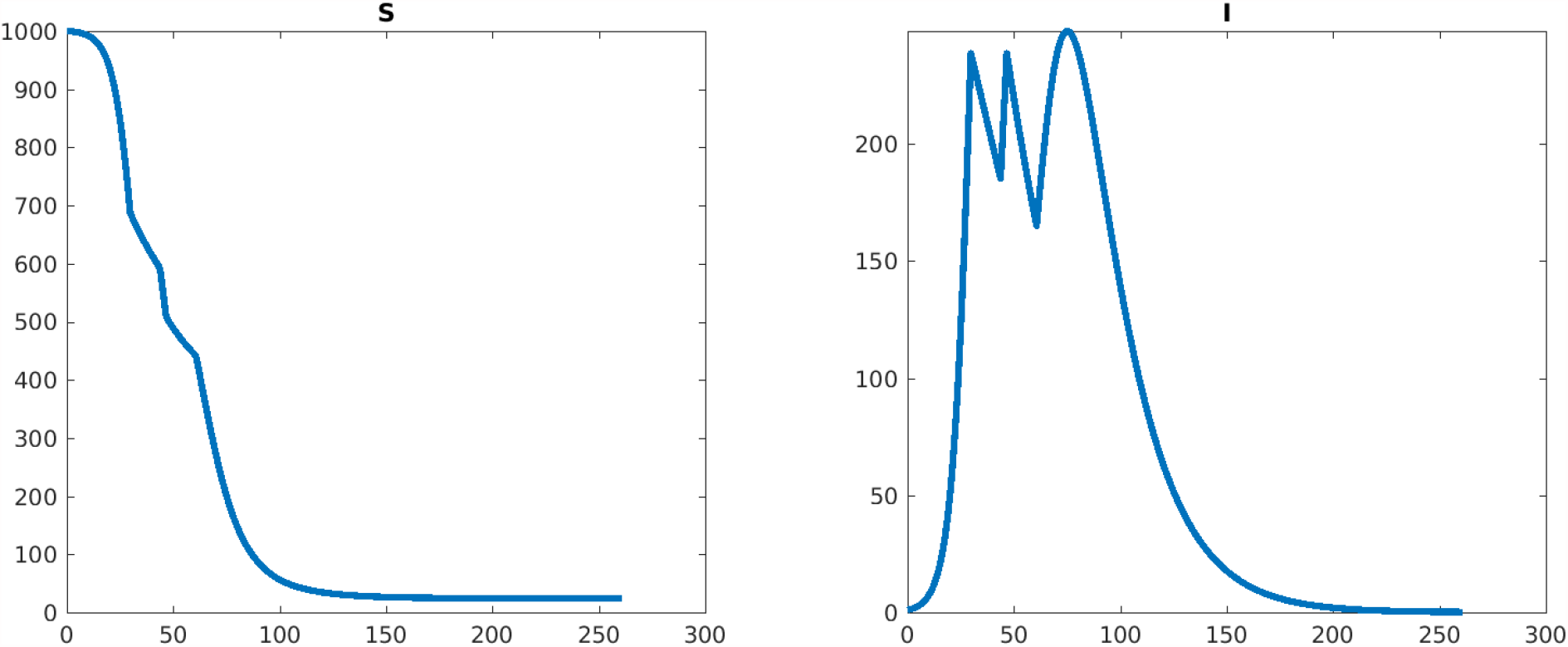
Plots of *S*(*t*) and *I*(*t*) (horizontal axis is time). Number of lockdowns: 2, peak *I* from formula: 238.740981, maximum of *I* on last (no lockdown) period: 248.153424. Lockdown start time(s): 29.73, 46.6.

**Figure 19:**
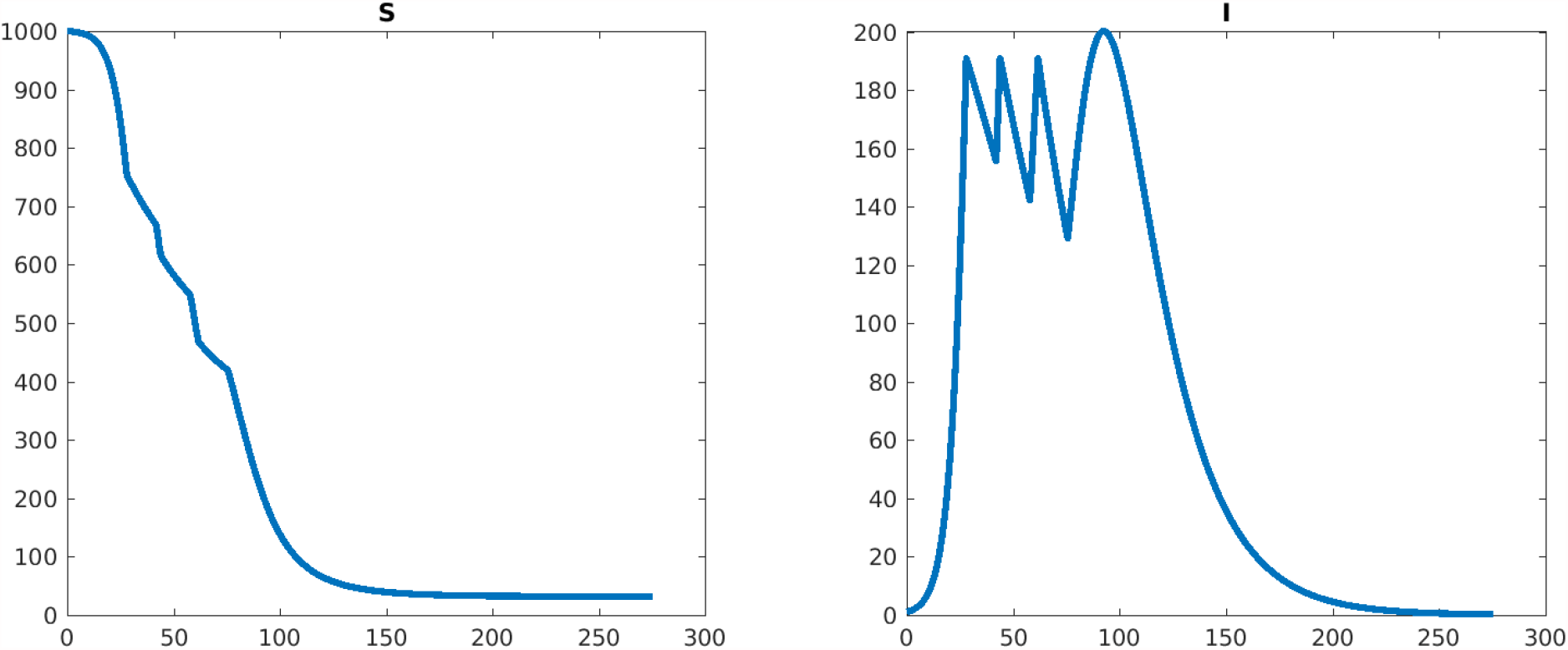
Plots of *S*(*t*) and *I*(*t*) (horizontal axis is time). Number of lockdowns: 3, peak *I* from formula: 190.862880, maximum of *I* on last (no lockdown) period: 200.218534. Lockdown start time(s): 28.01, 43.86, 61.6.

**Figure 20:**
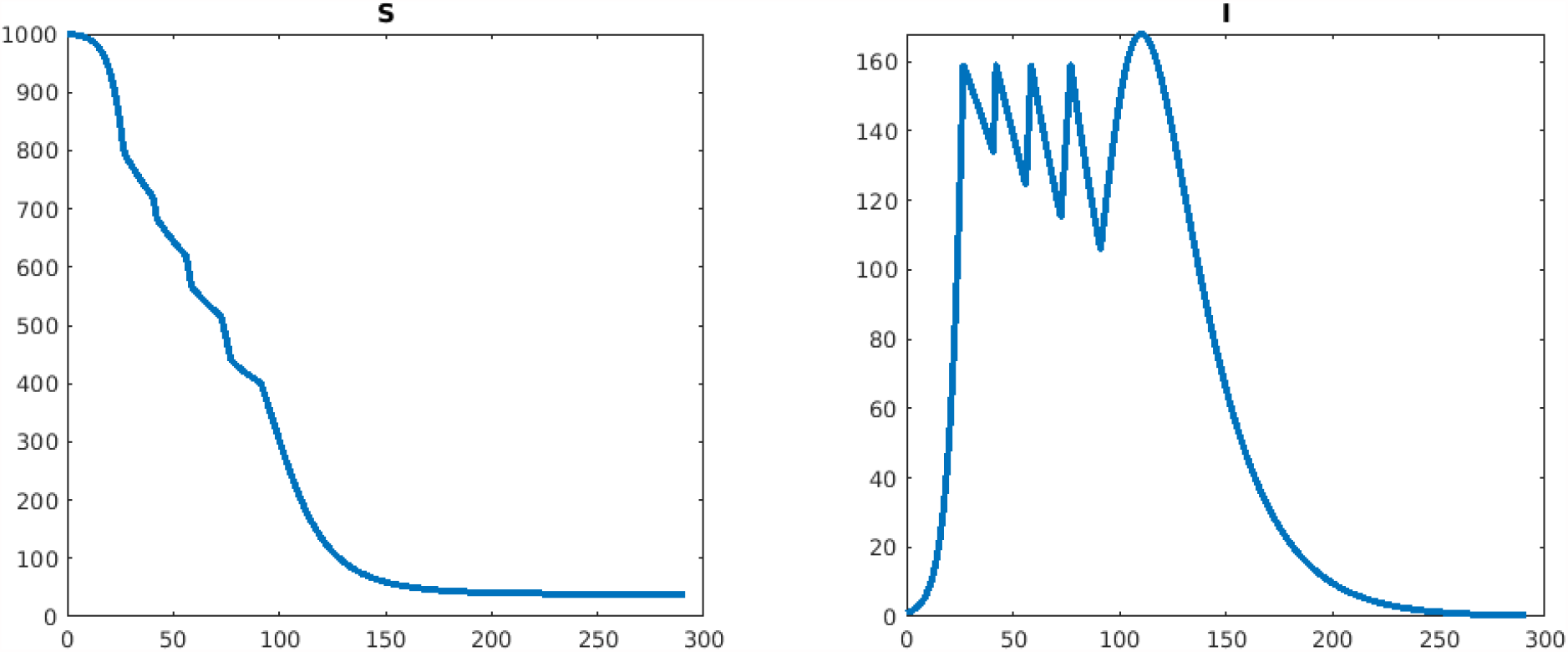
Plots of *S*(*t*) and *I*(*t*) (horizontal axis is time). Number of lockdowns: 4, peak *I* from formula: 158.980313, maximum of *I* on last (no lockdown) period: 167.977200. Lockdown start time(s): 26.74, 42.11, 58.60, 77.22.

**Figure 21:**
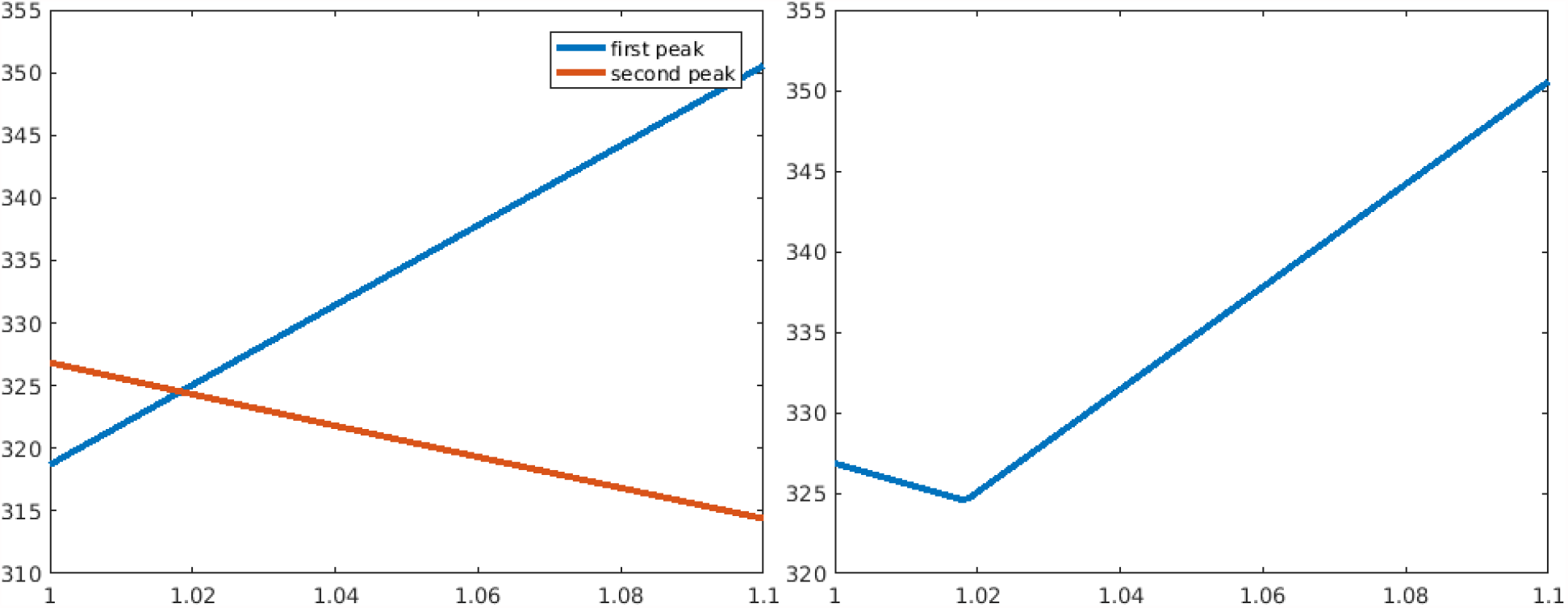
Left: Magnitude of first and second peak, with the lockdown time parametrized (*x* axis) by the percentage of the ideal (perfect lockdown) formula. ; 14 day lockdown. Note that the second peak decreases as the first peak happens later, so the minimum of the maximum among them will occur when the curves intersect. Right: The maximum between the curves; minimum is around fraction 1.018 of the optimal for perfect lockdowns, so trigger should occur later.

**Figure 22:**
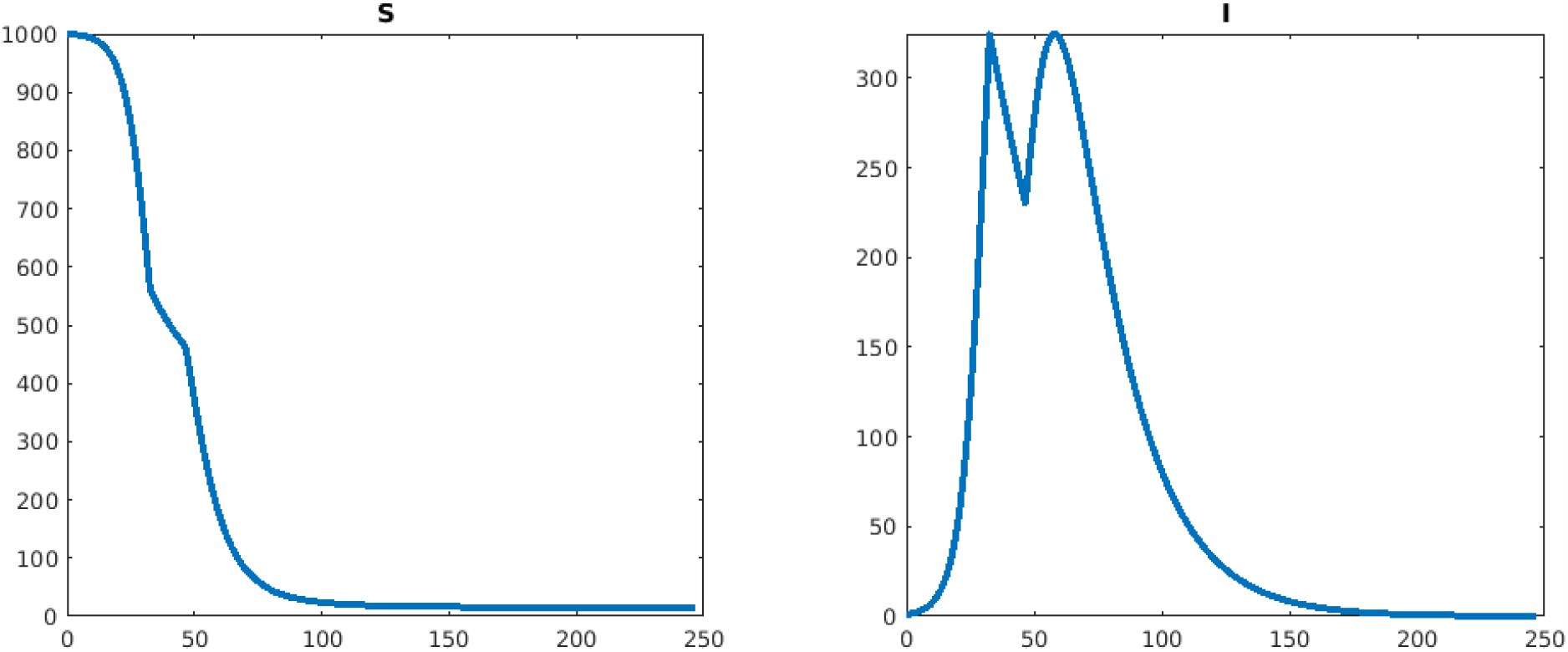
Simulation with an imperfect lockdown. Using now the optimal fraction 1.018 of the optimal for perfect lockdowns, obtained by minimizing the plot in Figure 21 (right). The optimal peak value is now approximately 324, which can be compared with the suboptimal plot (using the formula that assumed perfect lockdowns) shown in Figure 17, which had a peak of approximately 327. Observe how the two peaks are now balanced. The optimal result is not that different from the one obtained from our formulas, and the lockdown start is at time 30.39.

**Figure 23:**
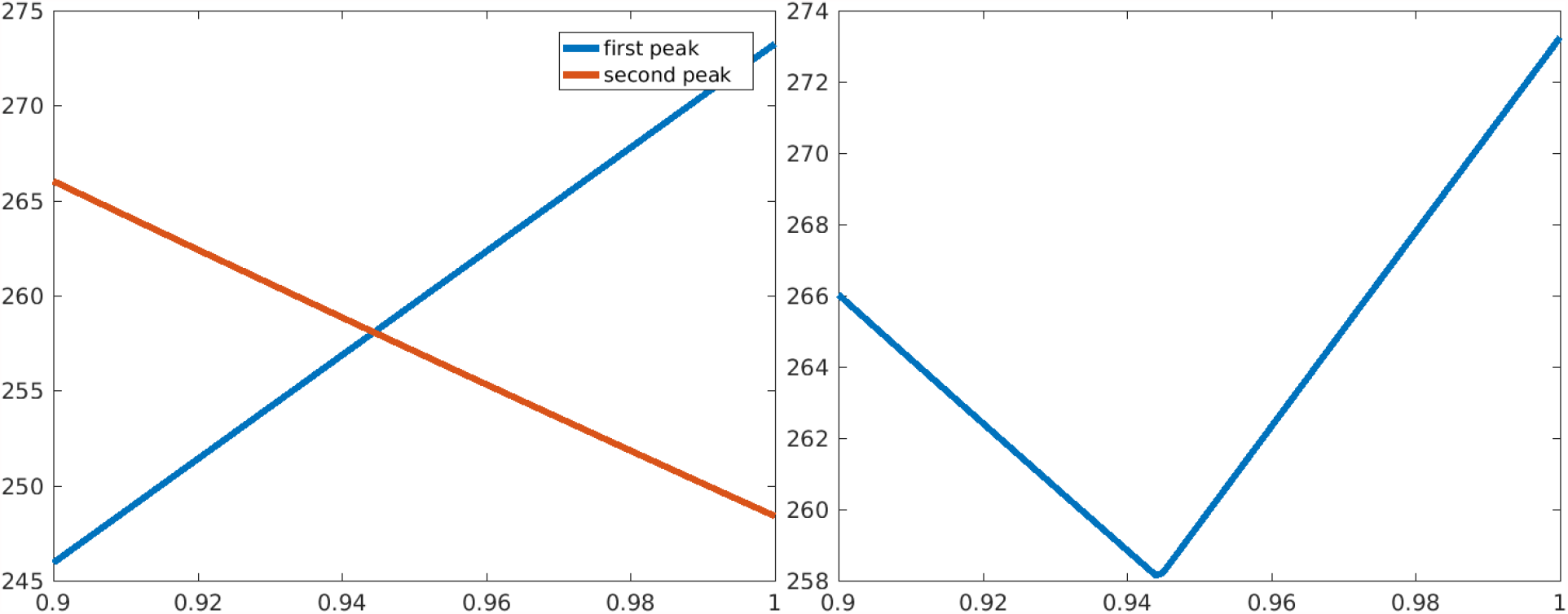
Left: Magnitude of first and second peak, with the lockdown time parametrized (*x* axis) by the percentage of the ideal (perfect lockdown) formula; 14 day lockdown. Note that the second peak decreases as the first peak happens later, so the minimum of the maximum among them will occur when the curves intersect. Right: The maximum between the curves; minimum is around fraction 0.944 of the optimal for perfect lockdowns, so trigger should occur earlier.

**Figure 24:**
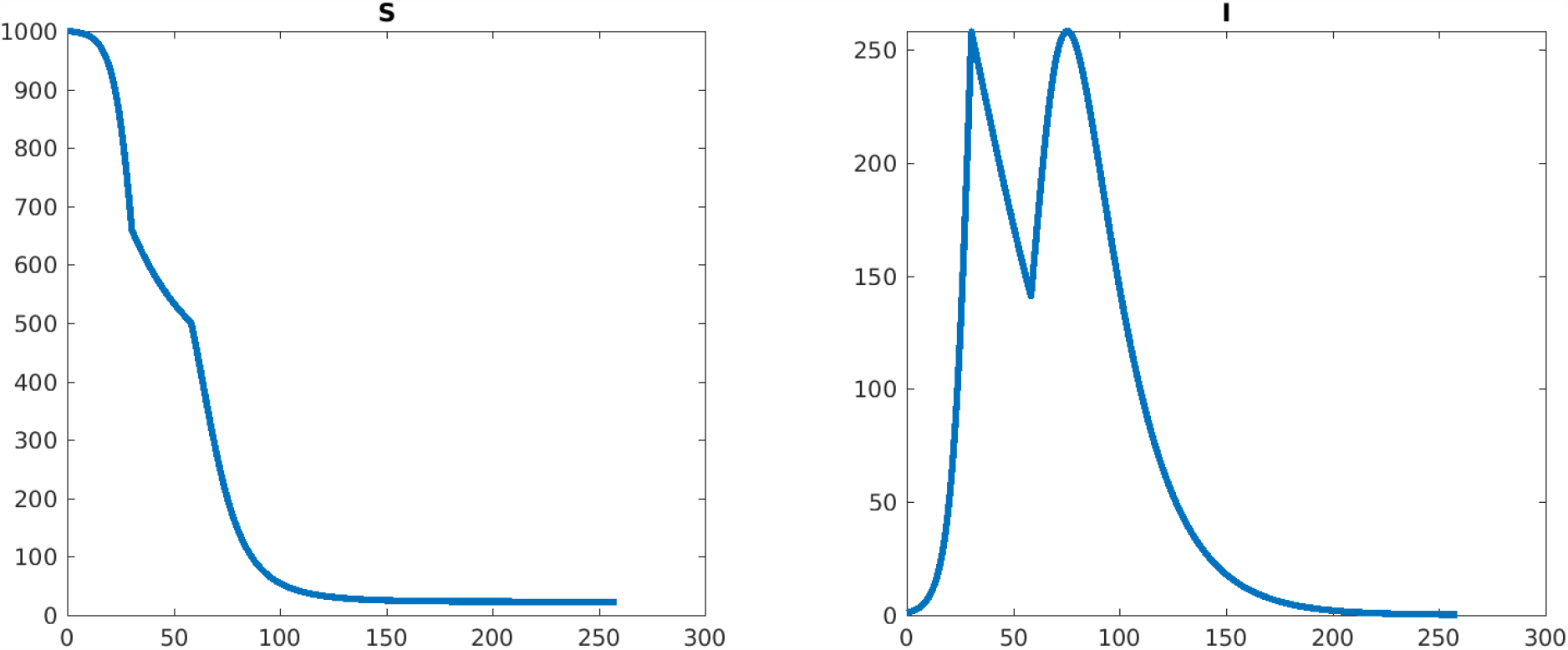
Simulation with an imperfect lockdown. Using now the optimal fraction 0.944 of the optimal for perfect lockdowns, obtained by minimizing the plot in Figure 23 (right). The optimal peak value is now approximately 258, which can be compared with the suboptimal plot (using the formula that assumed perfect lockdowns) shown in Figure 13, which had a peak of approximately 273. Observe how the two peaks are now balanced. The optimal result is very close to the one obtained from our formulas.

#### 4.4.2 Lockdown length is *T* = 14 days, *β*_0_ = 0.00005

In contrast, for 28-day lockdowns the plot in Figure 13 suggests that the formula derived for *β*_0_ = 0 waited too long for the first lockdown, so we first explore by what fraction < 1 to decrease the trigger point, from which the optimal strategy is clear.

## 5 Review of SIR model

To make this paper self-contained, we review here some facts about SIR models, see e.g. [19] or mathematical epidemiology texts for more details. We wish to analyze solutions, from initial conditions *S*(0) = *S*_0_, *I*(0) = *I*_0_.

### Infections always die-out in SIR model

Note that if *I*_0_ = 0, then *S*(*t*) ≡*S*_0_ and *I*(*t*) ≡ 0; in other words, every point of the form (*S*, 0) is an equilibrium. Similarly, if *S*_0_ = 0, then *S*(*t*) ≡0 and *I*(*t*) = *e*^−*νt*^*I*_0_ →0, so the case *S*_0_ = 0 is not interesting either. So we study the only interesting cases, *I*_0_ > 0 and *S*_0_ > 0.

Since 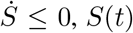 is a nonincreasing function of time, and thus *S*(*t*) ↘ *S*_∞_ for some *S*_∞_ ≥0. A most important result is this one:

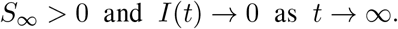

This says that the infection will end (asymptotically), and there will remain a number of “naive” individuals at the end.

We will show that *I*(*t*) → 0 and defer the proof that *S*_∞_ > 0 to later.

To prove this result, we will use this theorem: *if x*(*t*) *is a solution of a system of ODEs* 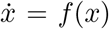, *and if the solution converges, x*(*t*) →*x*^*∗*^, *then x*^*∗*^ *must be an equilibrium point, i*.*e. f* (*x*^*∗*^) = 0. This is true because the omega-limit set of a trajectory is an invariant set, the LaSalle Invariance Principle (see e.g. [33]).

We apply this theorem as follows. First we define *V* (*t*) := *S*(*t*) + *I*(*t*), and notice that 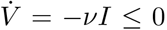, which means that *V* (*t*) is nonincreasing, and thus there is a limit *V* (*t*) ↘ *V*_∞_ as *t* → ∞. Therefore *I*(*t*) = *V* (*t*) − *S*(*t*) → *V*_∞_ − *S*_∞_ =: *I*_∞_ also has a limit. So the state *x*(*t*) = (*S*(*t*), *I*(*t*)) converges to *x*^*∗*^ := (*S*_∞_, *I*_∞_). It follows that *f* (*x*^*∗*^) = 0, which means that

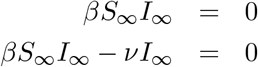

and from there we conclude that *I*_∞_ = 0 because *ν* ≠ 0.

We still have to show that *S*_∞_ > 0; we will in fact provide a formula for *S*_∞_.

### ℛ_0_ and epidemics

A central role in epidemiology is played by the “intrinsic reproductive rate”

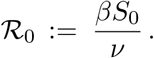

The epidemiological (and non-mathematically rigorous) definition of ℛ_0_ is “the average number of secondary cases produced by one infected individual introduced into a population of susceptible individuals,” where by a susceptible individual one means one who can acquire the disease. This can be made precise with a stochastic model, but an intuitive argument can be found in [19]. We remark ℛ_0_ has a generalization to more complex epidemics models, and it characterizes the local stability of the set of “disease-free” steady states (DFSS) (those for there are no infectives). One may compute ℛ_0_ using the so-called “next generation matrix” built from the differential equations. which was introduced in [34] (see e.g [35] and also the worked examples in [19]).

We discuss ℛ_0_ below in more detail, but for now note the following fact. From the ODE for *I*, we have that

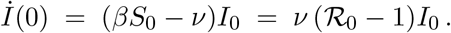

This means that *an epidemic will happen*, meaning that *I*(*t*) will increase when starting from any *I*(0) > 0, *if and only if* ℛ_0_ > 1.

Moreover, the initial growth of *I*(*t*) will be exponential, with rate *λ* = *ν* (ℛ_0_ −1). (For small times and a large susceptible population *S*_0_, we may assume that *S*(*t*) remains roughly constant.) Logarithmically plotting infections, we can estimate *λ*, and from there we may estimate

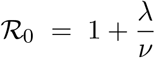

(assuming that we know *ν*, the recovery/death rate of infecteds), and

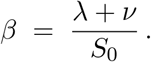

When ℛ_0_ ≤ 1 and *t* > 0,

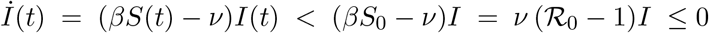

(because *S*(*t*) < *S*_0_) and so *I*(*t*) monotonically decreases to zero.

*From now on, when discussing the SIR model, we assume that* ℛ_0_ > 1.

### Peak infection time *t*_*p*_ and susceptibles at that time

The derivative *İ* = (*βS* − *ν*)*I* is positive for small *t*, because at zero it equals *ν* (ℛ_0_ − 1)*I* > 0.

On the other hand, since *I*(*t*) → 0 as *t* → ∞, the derivative must eventually become negative, which means that there is some time *t*_*p*_ (*p* for “peak” infectivity) at which *İ*(*t*_*p*_) = 0. Since *S*(*t*) decreases monotonically, the derivative of *I* can only change sign from positive to negative at exactly one such time *t*_*p*_. So *t*_*p*_ is the point at which *I*(*t*) attains is maximum.

From *İ* = 0 at *t*_*p*_, we have that

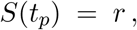

where we define for convenience

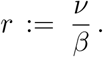

Thus, at the peak infection time, there is a precise formula for the number of susceptibles.

### A formula for the final number *S* _∞_ > 0 of susceptibles

Let us know derive an (implicit) equation for the limit *S*_∞_ of the susceptible population. We introduce the following function, along a given solution:

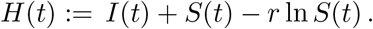

Taking derivatives,

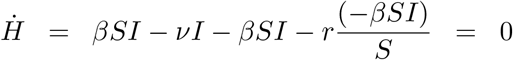

which means that *H* is constant along trajectories (a conserved quantity):

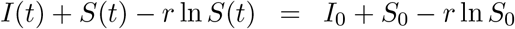

for all *t* > 0. It follows, in particular, that

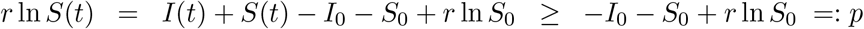

and therefore *S*(*t*) ≥ *e*^*pβ/ν*^ for all *t*, so taking limits *S*_∞_ ≥ *e*^*pβ/ν*^ > 0 as claimed.

Even better, we can obtain an equation for *S*_∞_ by passing to the limit in the conservation law, which gives (taking into account that *I*_∞_ = 0):

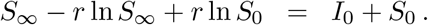

Dividing by *S*_0_ and using that ℛ_0_ = *βS*_0_*/ν*, and definining for convenience 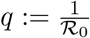, we obtain:

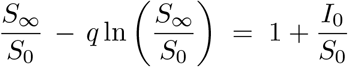

or, letting *x* := *S*_∞_*/S*_0_ and 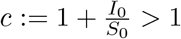:

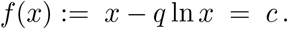

Observe that, since *S*(*t*) is decreasing, *x* < 1. We claim that there is exactly one solution of the equation *f* (*x*) = *c* with *x* ∈ (0, 1). By computing this solution, we can retrieve the final value of the susceptibles, *S*_∞_ = *xS*_0_. To prove that there is a solution *x* and it is unique, note that lim_*x→*0+_ *f* (*x*) = +∞ and *f* (1) = 1, and 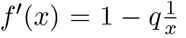 is an increasing function of *x*, with lim_*x→*0+_ *f ′* (*x*) = −∞and *f ′*(1) = 1 −*q* > 0 (because we assumed ℛ_0_ > 1). Therefore, *f* decreases until some *x*^*∗*^ and then increases back to 1. Since *c* > 1, it follows that *f* (*x*) = *c* has a unique solution, as we wanted to prove.

There is in fact a solution of this equation that employs a classical function. For simplicity let us write *s* := *q*^−1^ = ℛ_0_. Multiplying by −*s*, we write the equation as ln *x* −*sx* = −*sc*. Taking exponentials and multiplying again by −*s* results in *we*^*w*^ = *y*, where *w* := −*sx* and *y* := −*se*^−*sc*^. Note that, since *s* > 1 and *c* > 1, *y* ∈ (− 1*/e*, 0). The function *w ↦ we*^*w*^ has an inverse, defined on (− 1*/e*, 0), called the *Lambert W function* (MATLAB command `lambertw`). So, *w* = *W* (*y*), and since *x* = −*w/s*, we conclude that

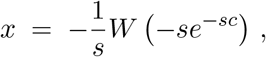

or, after multiplying by *S*_0_:

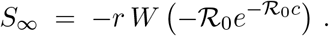

Typically, *I*_0_ ≈ 0 (one individual is enough to cause an epidemic), so *c* ≈ 1 and in that case

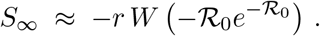

If one can measure the proportion of people who did not get sick compared to the total initial population, then one can solve for ℛ_0_. This is one way to compute ℛ_0_ from historical data.

### A formula for the peak value *I*(*t*_*p*_) of infectives

Determining the peak value *I*(*t*_*p*_) is of critical importance in practice. If a proportion *θ* of infected individuals will need hospital care, one can then predict, early on in an infection (and assuming that the SIR model is correct), the maximum number *θI*(*t*_*p*_) of people who will require hospital beds (or intensive care treatment) at any given time, and thus enforce a more stringent NPI policy if this number is projected to overwhelm hospital capacity.

Let us take again the conservation law

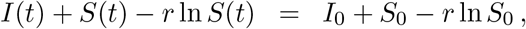

and now specialize at *t* = *t*_*p*_, using that *S*(*t*_*p*_) = *r*. Then,

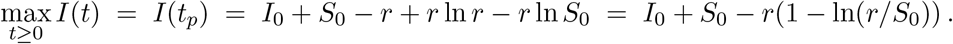

Another way to write this is to use that *r/S*_0_ = 1*/*ℛ_0_, so

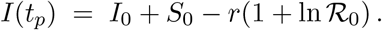

#### 5.0.1 Alternative definitions of ℛ_0_

There are alternative definitions of ℛ_0_ (for the SIR model) t hat one often encounters in the literature: ℛ_0_ = *βN/ν*, where *N* is the total population size, or even ℛ_0_ = *β/ν*. Let us quickly explain how these relate to what we are doing here. As alluded to earlier, the general definition of ℛ_0_ is given in terms of what is called a “disease-free steady state” (DFSS), meaning a steady state in which there are no infected individuals. For the specific case of the SIR model, this would mean any steady state of the form *S* = *S*_0_, *I* = 0, and *R* = *N* −*S*_0_. With this definition, ℛ_0_ = *βS*_0_*/ν*, but there are many possible ℛ_0_’s depending on what is the number or removed individuals at the initial time. In particular, for the equilibrium with *R* = 0, ℛ_0_ = *βN/ν*.

What about the definition ℛ _0_ = *β/ν*? It is often the case that one normalizes the population to fractions: *S*_*f*_ := *S/N, I*_*f*_ := *I/N*, 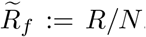. In this case, 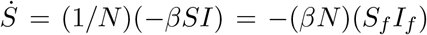 and *İ*_*f*_ = (1*/N*)(*βS* − *ν*)*I* = ((*βN*)*S*_*f*_ − *ν*)*I*_*f*_, so

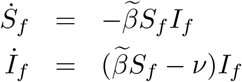

with 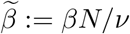. Now 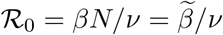 in terms of this new *β*. Note that *S*_*f*_ and *I*_*f*_ are dimensionless and that 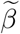 has units of (1/time), while the original *β* had units of 1/(time × individuals), so 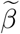 is perhaps more elegant.

We prefer not to perform this normalization because, when there are “vital dynamics” such as immigration, emigration, births, and/or deaths, the total population *N* would not be constant.

## 6 Discussion

In this paper we discussed an SIR model with strict (no-contact) lockdowns. We studied the problem of deciding when to start each one of *K* of lockdowns, with respective lengths *T*_*k*_, *k* = 1, …, *K*, so as to minimize the maximum number of infected individuals at any given time, and provided an exact formula in which lockdowns should start whenever the number of infectious individuals reaches 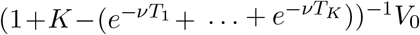, where *V*_0_ is the “virtual peak” that would have resulted from no lockdowns.

As we discussed, a perfect or near-perfect lockdown is not practical, but the ideal case helps understand the problem with non-strict lockdowns, and we presented numerical evidence that the formula is reasonable accurate even in the not totally (but still reasonably) strict case. The fellow-up paper [36] has recently started the study of the same optimization problem in the non-strict case, showing that in the optimal case the subsequent peak after release from one lockdown coincides with the infective population at the start of the lockdown. Much further work remains, including the extension of the perfect-lockdown problem to models such as the ones presented in [20, 25, 26].

We also presented results showing that equal-length lockdowns are optimal, and we quantified the sensitivity to poorly known contagion rates.

## Data Availability

No data is used.

## 7 Acknowledgments

We are grateful to James Greene for many discussions, and to an anonymous referee for the suggestion of the equal-lengths result. Parts of this work were supported by grants NSF 1849588, ONR N00014-21-1-2431, and AFOSR FA9550-21-1-0289.

